# Bringing Critical Race Praxis into the Study of Electrophysiological Substrate of Sudden Cardiac Death: The Atherosclerosis Risk in Communities (ARIC) Study

**DOI:** 10.1101/19009167

**Authors:** Kelly Jensen, Stacey J. Howell, Francis Phan, Maedeh Khayyat-Kholghi, Linda Wang, Kazi T. Haq, John Johnson, Larisa G. Tereshchenko

**Author notes:** Correspondence: Larisa Tereshchenko, 3181 SW Sam Jackson Park Rd; UHN62; Portland, OR, 97239.

## Abstract

Race is an established risk factor for sudden cardiac death (SCD). We sought to determine whether the association of electrophysiological (EP) substrate with SCD varies between black and white individuals. Participants from the Atherosclerosis Risk in Communities study with analyzable ECGs (n=14,408; age 54±6 y; 74% white) were included. EP substrate was characterized by traditional 12-lead ECG and vectorcardiographic metrics. Two competing outcomes were adjudicated SCD and non-sudden cardiac death (nonSCD). Interaction of ECG metrics with race was studied in Cox proportional hazards and Fine-Gray competing risk models, adjusted for prevalent cardiovascular disease (CVD), risk factors, and incident non-fatal CVD. At the baseline visit linear regression analysis, adjusted for age, sex, and study center, showed black individuals had larger Spatial Ventricular Gradient magnitude by 0.30 (95%CI 0.25-0.34) mV, SAI QRST by 18.4 (13.7-23.0) mV*ms, Cornell voltage by 0.30 (95%CI 0.25-0.35) mV than white individuals. Over a median follow-up of 24.4 years, SCD incidence was higher in black (2.86; 95%CI 2.50-3.28 per 1000 person-years) than white individuals (1.37; 95%CI 1.22-1.53 per 1000 person-years). Black individuals with hypertension had the highest rate of SCD: 4.26; 95%CI 3.66-4.96 per 1000 person-years. Race did not modify associations of EP substrate with SCD and nonSCD. EP substrate does not explain racial disparities in SCD rate.

## Introduction

Sudden cardiac death (SCD) disproportionately affects black individuals, with a nearly two-fold increased risk of SCD in black as compared to white individuals.^1-3^ Socioeconomic disparities and the burden of cardiovascular (CV) risk have been proposed as factors that may underlie the observed racial differences in SCD.^1^ However, adjustment for socioeconomic and behavioral measures of health does not fully explain an excess of SCD risk in black individuals.^1, 2, 4^ Moreover, the large prospective community-based cohort the Atherosclerosis Risk in Communities (ARIC) study showed that race modifies an association of several major risk factors with SCD. Prevalent coronary heart disease (CHD) and body mass index (BMI) carried greater risk of SCD in white as compared to black individuals.^1^ In contrast, hypertension carried significantly larger risk of SCD in black as compared to white individuals.^1^ The mechanisms behind these observed interactions remain unclear but pose the question of differences in EP substrate metrics between the two racial groups.

A conventional 12-lead electrocardiogram (ECG) characterizes global electrophysiological (EP) substrate of SCD,^5^ which can be assessed by traditional [QRS duration, QTc interval, ECG-left ventricular hypertrophy (LVH)] and novel metrics, such as global electrical heterogeneity (GEH).^6, 7^ GEH is measured by spatial QRS-T angle, magnitude and direction (elevation and azimuth) of the spatial ventricular gradient (SVG) vector, and its scalar value sum absolute QRST integral (SAI QRST). The addition of GEH to known CV risk factors improves reclassification of SCD.^8^ The SVG vector describes the magnitude and direction of the steepest gradient between the areas of the heart with the longest and the shortest total recovery time.^9-11^ The SVG is related to global heterogeneity of action potential duration and morphology^12, 13^ through the heart. While racial differences in ECG-LVH diagnostic criteria are recognized^14^, it is unknown whether there are racial differences in association between EP substrate and risk of SCD.

The self-identified race is a social construct. Recently, the Public Health Critical Race Praxis^15^ was developed as a theoretical framework for studies of racial disparities. Our goal was to apply the Public Health Critical Race Praxis^15^ approach to the epidemiological study of SCD substrate. We hypothesized that (1) there are racial differences in global ECG measures of EP substrate, and (2) self-identified race modifies the association of EP substrate with SCD.

## Methods

The ARIC Study data are available through the National Heart, Lung, and Blood Institute’s Biological Specimen and Data Repository Information Coordinating Center (BioLINCC)^16^ and the National Center of Biotechnology Information’s database of Genotypes and Phenotypes (dbGaP).^17^

### Study population

The Atherosclerosis Risk in Communities (ARIC) study recruited 15,792 participants (age 45-64 y) in 1987-1989. All participants underwent standardized examinations.^18^ In this study, we included ARIC cohort participants with recorded resting 12-lead ECG and measured global electrical heterogeneity (GEH)^8^; n=15,777. We excluded participants who self-identified themselves as non-white or non-black race (n=48), or as black at the Washington County, and Minneapolis field centers (n=55), those with missing covariates (n=1,220), outcome (n=1), and non-sinus median beat (n=45). The final sample of participants with normal sinus median beat included 14,408 participants.

### Exposures of race and electrocardiographic global electrical heterogeneity

The race was self-reported. We analyzed resting 12-lead ECGs of the first five study visits. Visit 1 was conducted in 1987-1989, visit 2 in 1990-1992, visit 3 in 1993-1995, visit 4 in 1996-1998, and visit 5 in 2011-2013. Traditional ECG amplitudes and intervals were measured by the 12 SL algorithm as implemented in the Magellan ECG Research Workstation V2 (GE Marquette Electronics, Milwaukee, WI), and Cornell voltage was calculated.

GEH was measured by spatial QRS-T angle, SVG magnitude, azimuth, and elevation, and SAI QRST.^19^ Both area and peak SVG vectors and QRS-T angles were measured.^19^ We used normal sinus time-coherent median beat with identified isoelectric heart vector origin point.^20^ The MATLAB (MathWorks, Natick, MA, USA) open-source software code for GEH measurement and the heart vector origin definition is provided at https://physionet.org/physiotools/geh and https://github.com/Tereshchenkolab/Origin.

### Primary outcome: sudden cardiac death

Follow-up of ARIC participants^21^ and determination of SCD has been described in prior reports.^22^ SCD, the primary study outcome, was defined as a sudden pulseless condition in a previously stable participant without a non-cardiac cause of arrest if it occurred outside of the hospital or in the emergency department. It was classified as definite, probable, or possible based on physician-adjudication.

### Competing mortality outcome: non-sudden cardiac death

Competing non-sudden cardiac death (nonSCD) was defined as an SCD exclusion, a composite of fatal CHD, heart failure (HF) death, death in a participant with baseline HF, or incident hospitalized HF. Fatal CHD cases were adjudicated by the ARIC Morbidity and Mortality Classification Committee.^21, 23^ Baseline prevalent HF was based on Gothenburg criteria (stage 3 symptomatic HF with both cardiac and pulmonary symptoms and current medical treatment^24^), or self-reported use of HF medication. Incident HF was defined as presence of HF codes in a death certificate or an International Classification of Diseases (*ICD-*9) discharge code, as previously described.^25^ All other deaths were included in the noncardiac death outcome.

### Baseline clinical characteristics

Body mass index (BMI) was categorized as underweight (<18.5 kg/m^2^), normal weight (18.5 to <25.0 kg/m^2^), overweight (25.0 to <30.0 kg/m^2^) or obese (≥30.0 kg/m^2^). Hypertension was defined as blood pressure (BP) of ≥140/90 mm Hg, or self-reported antihypertensive medications at visit 1. Diabetes was defined as nonfasting blood glucose ≥200 mg/dL, fasting blood glucose ≥126 mg/dL, self-reported physician diagnosis of diabetes, or self-reported medications for diabetes or high blood sugar at visit 1. Stages of chronic kidney disease (CKD) were based on estimated glomerular filtration rate (eGFR) calculated using the CKD Epidemiology Collaboration equation (CKD-EPI).^26^ Participants were classified into stage 1 CKD (eGFR_CKD-EPI_ ≥90 mL/min/1.73 m^2^), stage 2 CKD (eGFR_CKD-EPI_ 60 to <90 mL/min/1.73 m^2^), Stage 3 CKD (eGFR_CKD-EPI_ 30 to <60 mL/min/1.73 m^2^), stage 4 CKD (eGFR_CKD-EPI_ 15 to <30 mL/min/1.73 m^2^), and stage 5 CKD (eGFR_CKD-EPI_ <15 mL/min/1.73 m^2^) or established kidney failure. Baseline serum electrolytes concentrations were measured in the central laboratory.

Prevalent stroke was based on a previously reported stroke and transient ischemic attack diagnostic algorithm^27^. Prevalent CHD included a self-reported physician diagnosis of myocardial infarction (MI), baseline ECG evidence of MI by the Minnesota code^28^, or a history of coronary revascularization (either via coronary artery bypass surgery or percutaneous coronary intervention). The use of antiarrhythmic drugs included self-reported and validated by medications inventory use of class I, II (beta-blockers), III, IV (phenylalkylamines, and benzothiazepines calcium channel blockers), or V (digoxin) antiarrhythmic agents.

### Socioeconomic factors

Socioeconomic status was assessed during a home interview at visit 1. Total combined family income for the past 12 months (in 1987-1989 values) was self-reported in one of the following categories: under $5,000; $5,000-$7,999; $8,000-$11,999; $12,000-$15,999; $16,000-$24,999; $25,000-$34,999; $35,999-$49,999; over $50,000, or not reported. Lifetime educational level was defined as the highest grade or year of school completed. The most recent occupation was recorded in one of the following categories: (1) Managerial and Professional Specialty Occupations; (2) Technical, Sales, and Administrative Support Occupations; (3) Service Occupations; (4) Farming, Forestry, and Fishing Occupations; (5) Precision Production, Craft, and Repair Occupations; (6) Operators, Fabricators, and Laborers; (7) Homemakers; (8) Retired; (9) Others.

Baseline physical activity was measured at work, in sport, and during leisure time, using the modified Baecke questionnaire^29^, which defined semicontinuous indices ranging from 1 (low) to 5 (high). Current cigarette smoking and consumption of alcoholic beverages was used as self-reported at visit 1. Self-reported health insurance status was ascertained at visit 1.

### Incident non-fatal cardiovascular events

Incident non-fatal CV events included atrial fibrillation (AF), stroke, CHD, and HF. Incident AF included AF detected on follow-up 12-lead ECG or hospital discharge records (*ICD-9* code 427.3).^30^ Physician-adjudicated definite or probable incident strokes were included.^31^ Incident CHD was physician-adjudicated and included definite or probable MI or a coronary revascularization procedure.^21, 23^ Incident HF was defined above.^25^

### Statistical analyses

#### Cross-sectional analyses of visit 1 data

To investigate differences in global ECG metrics between black and white individuals, we performed cross-sectional linear regression analyses using visit 1 data. Model 1 was adjusted for age, sex, and study center. To determine whether racial differences in GEH could be explained by racial disparities, including differences in socioeconomic, traditional and novel clinical risk factors, Model 2 was additionally adjusted for prevalent cardiovascular (CV) disease (HF, CHD, stroke), known CV risk factors (diabetes, hypertension, current smoking and alcohol intake, work, sport, and leisure physical activity levels, levels of total cholesterol, high density lipoprotein, and triglycerides, BMI), use of antihypertensive and antiarrhythmic medications, serum concentrations of sodium, potassium, calcium, magnesium, phosphorus, and uric acid, total protein and albumin, blood urea nitrogen, CKD stage classified by eGFR_CKD-EPI_, traditional ECG characteristics (mean heart rate, QRS duration, Bazett-corrected QT interval, Cornell voltage), and socioeconomic factors (education level, occupation category, income, health insurance).

#### Analysis of circular variables

Circular variables were analyzed as previously described.^32^ SVG azimuth was transformed by doubling its value and then adding 360°.^33^

#### Survival analyses

We built Cox proportional hazards and Fine-Gray competing risks models. The proportional-hazards assumption was verified using *stcox PH-assumptions* suite of tests implemented in STATA (StataCorp LP, College Station, TX) for all predictors of interest in most models.

Exceptions were stated. To standardize comparisons, all continuous ECG variables were expressed as by their z-score. To adjust for confounders, we constructed two models, performed a statistical test for interaction with race in each model, and constructed race-stratified models for white and black individuals. Relative hazard ratio (RHR) with a 95% confidence interval (CI) of SCD risk for black relative to white individuals was reported, assuming HR for white individuals is a reference.

Model 1 was adjusted for demographic characteristics (age, sex, and study center), prevalent CV disease (HF, CHD, stroke), baseline CV risk factors (diabetes, hypertension, levels of total cholesterol, high density lipoprotein, and triglycerides, BMI, use of antihypertensive and antiarrhythmic medications, serum concentrations of sodium, potassium, calcium, magnesium, phosphorus, and uric acid, total protein and albumin, blood urea nitrogen, CKD stage classified by eGFR_CKD-EPI_,), and measured at visit 1 socioeconomic factors (smoking and alcohol intake, work, sport, and leisure physical activity levels, education level, occupation category, income, health insurance). Time-updated model 2 included time-updated ECG predictors (one-by-one), all baseline covariates included in model 1, and time-updated incident nonfatal CV events (AF, HF, CHD, and stroke). Race-stratified associations of continuous ECG variables with SCD were also studied using adjusted (model 1) Cox regression models incorporating cubic splines with 4 knots.

To compare competing risks of SCD and nonSCD, we constructed two Fine and Gray’s competing risks models^34^ for SCD and nonSCD outcomes, using the same covariates as in Cox models. We calculated the relative sub-hazard ratio (RSHR) with 95% CI of SCD risk for black relative to white individuals, assuming SHR for white participant is a reference.

Based on a recent study demonstrating a significantly stronger association of hypertension with SCD in black as compared to white individuals,^1^ we additionally constructed a set of models that included interaction with both race and hypertension categories. The white hypertension-free subgroup was the reference.

Statistical analyses were performed using STATA MP 16.1 (StataCorp LP, College Station, TX). Considering the many multivariate analyses performed, statistical significance at the 0.05 level should be interpreted cautiously.

## Results

### Study population

Black study participants (Table 1) were slightly younger, with a higher prevalence of HF, stroke, and major risk factors (hypertension, diabetes, smoking) than white individuals. White participants had a higher prevalence of CHD, higher level of triglycerides, were less physically active at work, but more physically active at leisure and sport than black participants. There were large disparities between races in access to healthcare, income, education, and occupation.

**Table 1.**
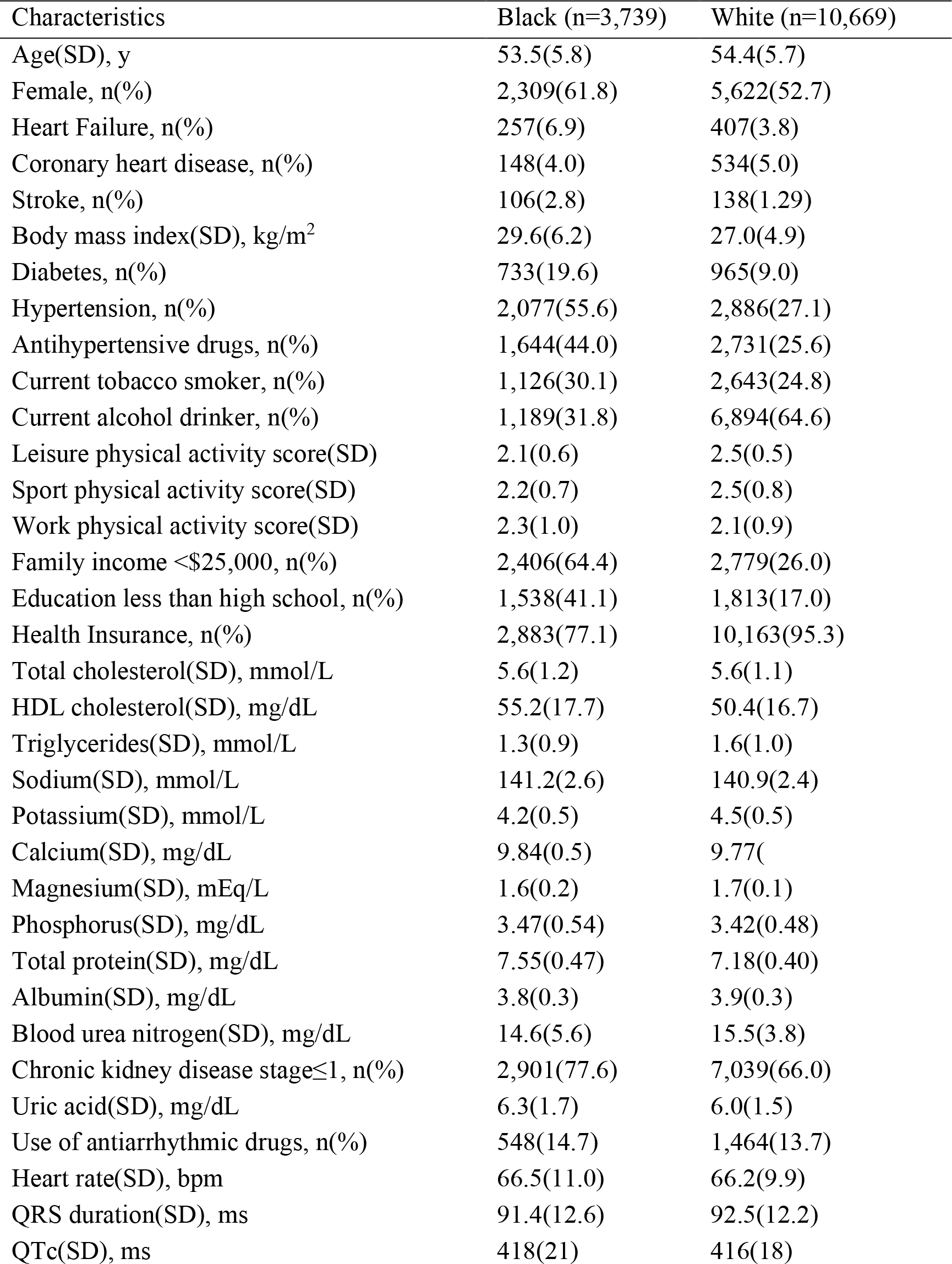
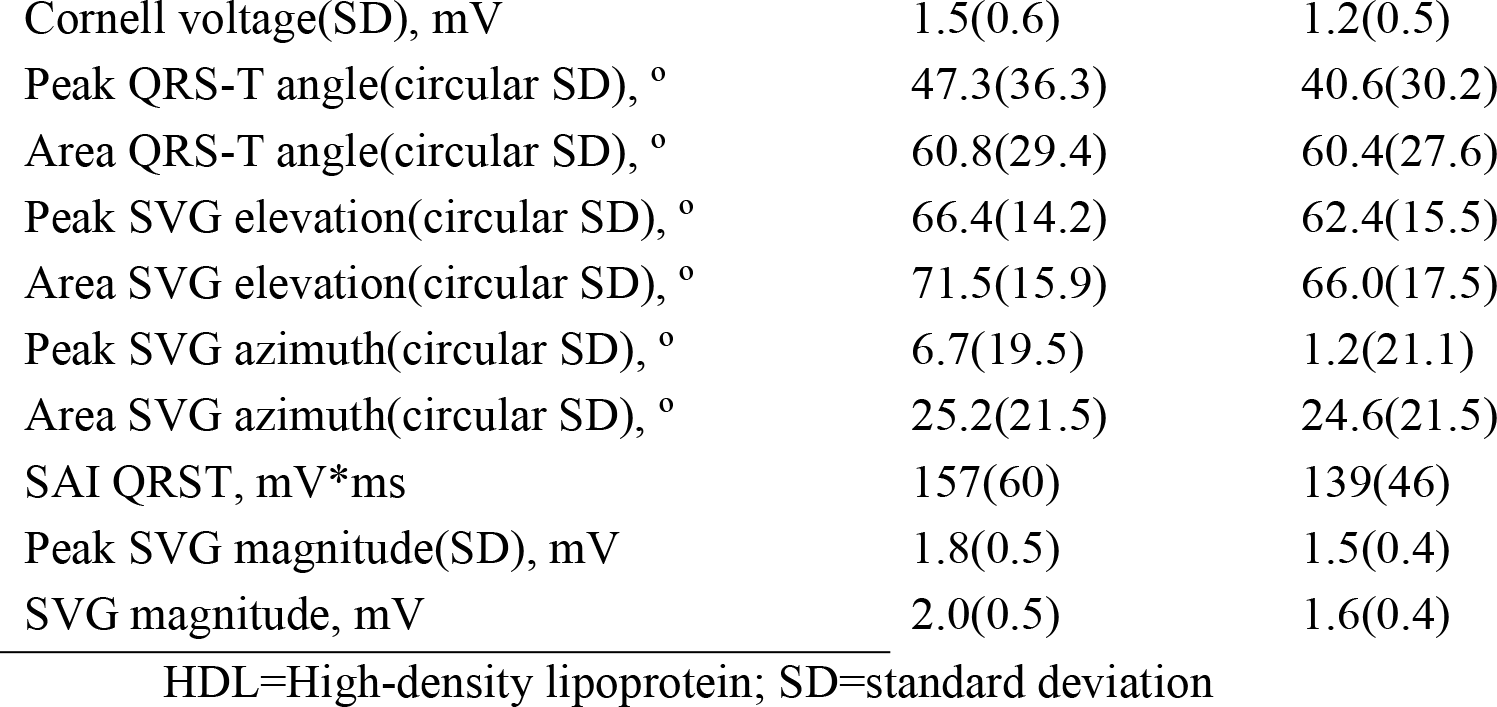
Comparison of baseline clinical and ECG characteristics in black and white participants

| Characteristics | Black (n=3,739) | White (n=10,669) |
| --- | --- | --- |
| Age(SD), y | 53.5(5.8) | 54.4(5.7) |
| Female, n(%) | 2,309(61.8) | 5,622(52.7) |
| Heart Failure, n(%) | 257(6.9) | 407(3.8) |
| Coronary heart disease, n(%) | 148(4.0) | 534(5.0) |
| Stroke, n(%) | 106(2.8) | 138(1.29) |
| Body mass index(SD), kg/m<sup>2</sup> | 29.6(6.2) | 27.0(4.9) |
| Diabetes, n(%) | 733(19.6) | 965(9.0) |
| Hypertension, n(%) | 2,077(55.6) | 2,886(27.1) |
| Antihypertensive drugs, n(%) | 1,644(44.0) | 2,731(25.6) |
| Current tobacco smoker, n(%) | 1,126(30.1) | 2,643(24.8) |
| Current alcohol drinker, n(%) | 1,189(31.8) | 6,894(64.6) |
| Leisure physical activity score(SD) | 2.1(0.6) | 2.5(0.5) |
| Sport physical activity score(SD) | 2.2(0.7) | 2.5(0.8) |
| Work physical activity score(SD) | 2.3(1.0) | 2.1(0.9) |
| Family income <\$25,000, n(%) | 2,406(64.4) | 2,779(26.0) |
| Education less than high school, n(%) | 1,538(41.1) | 1,813(17.0) |
| Health Insurance, n(%) | 2,883(77.1) | 10,163(95.3) |
| Total cholesterol(SD), mmol/L | 5.6(1.2) | 5.6(1.1) |
| HDL cholesterol(SD), mg/dL | 55.2(17.7) | 50.4(16.7) |
| Triglycerides(SD), mmol/L | 1.3(0.9) | 1.6(1.0) |
| Sodium(SD), mmol/L | 141.2(2.6) | 140.9(2.4) |
| Potassium(SD), mmol/L | 4.2(0.5) | 4.5(0.5) |
| Calcium(SD), mg/dL | 9.84(0.5) | 9.77( |
| Magnesium(SD), mEq/L | 1.6(0.2) | 1.7(0.1) |
| Phosphorus(SD), mg/dL | 3.47(0.54) | 3.42(0.48) |
| Total protein(SD), mg/dL | 7.55(0.47) | 7.18(0.40) |
| Albumin(SD), mg/dL | 3.8(0.3) | 3.9(0.3) |
| Blood urea nitrogen(SD), mg/dL | 14.6(5.6) | 15.5(3.8) |
| Chronic kidney disease stage≤1, n(%) | 2,901(77.6) | 7,039(66.0) |
| Uric acid(SD), mg/dL | 6.3(1.7) | 6.0(1.5) |
| Use of antiarrhythmic drugs, n(%) | 548(14.7) | 1,464(13.7) |
| Heart rate(SD), bpm | 66.5(11.0) | 66.2(9.9) |
| QRS duration(SD), ms | 91.4(12.6) | 92.5(12.2) |
| QTc(SD), ms | 418(21) | 416(18) |
| Cornell voltage(SD), mV | 1.5(0.6) | 1.2(0.5) |
| Peak QRS-T angle(circular SD), ° | 47.3(36.3) | 40.6(30.2) |
| Area QRS-T angle(circular SD), ° | 60.8(29.4) | 60.4(27.6) |
| Peak SVG elevation(circular SD), ° | 66.4(14.2) | 62.4(15.5) |
| Area SVG elevation(circular SD), ° | 71.5(15.9) | 66.0(17.5) |
| Peak SVG azimuth(circular SD), ° | 6.7(19.5) | 1.2(21.1) |
| Area SVG azimuth(circular SD), ° | 25.2(21.5) | 24.6(21.5) |
| SAI QRST, mV*ms | 157(60) | 139(46) |
| Peak SVG magnitude(SD), mV | 1.8(0.5) | 1.5(0.4) |
| SVG magnitude, mV | 2.0(0.5) | 1.6(0.4) |

HDL=High-density lipoprotein; SD=standard deviation

### Differences in ECG metrics between black and white participants

SVG magnitude, SAI QRST, and Cornell voltage were significantly larger in black than white individuals, even after adjustment for confounders (Table 2 and Figure 1). The spatial QRS-T angle and direction of SVG, as well as QTc were similar in white and black individuals (Supplemental Figure 1).

**Table 2.** Difference in GEH variables in black as compared to white participants

| GEH characteristic | Model 1 | Model 2 |
| --- | --- | --- |
|  | Difference (95%CI) | Difference (95%CI) |
| Peak QRS-T angle, ° | +11(8.6-15.0) | +5.2(2.2-8.3) |
| Area QRS-T angle, ° | +1.4(-1.2 to 4.1) | -2.7(-5.2 to -0.3) |
| Peak SVG elevation, ° | +4.6(3.1-6.0) | +0.8(-0.5 to 2.2) |
| Area SVG elevation, ° | +6.1(4.5-7.8) | +2.3(0.6-3.9) |
| Peak SVG azimuth, ° | +11.0(5.8-15.6) | +0.3(-4.5 to 5.0) |
| Area SVG azimuth, ° | +1.8(-3.1 to 6.7) | -11.3(-15.9 to -6.6) |
| SAI QRST, mV*ms | +18.4(13.7-23.0) | +14.0(9.8-18.0) |
| Peak SVG magnitude, mV | +0.26(0.21-0.30) | +0.22(0.18-0.26) |
| SVG magnitude, mV | +0.30(0.25-0.34) | +0.28(0.23-0.33) |
| Cornell voltage, mV | +0.30(0.25-0.35) | +0.22(0.18-0.27) |
| QRS duration, ms | -0.4(-1.6 to 0.7) | -3.3(-4.4 to -2.2) |
| QTc, ms | +2.4(0.5-4.2) | -0.6(-2.3 to 1.1) |
| Heart rate, bpm | -0.8(-1.8 to 0.2) | -3.2(-4.2 to -2.3) |

**Figure 1.**
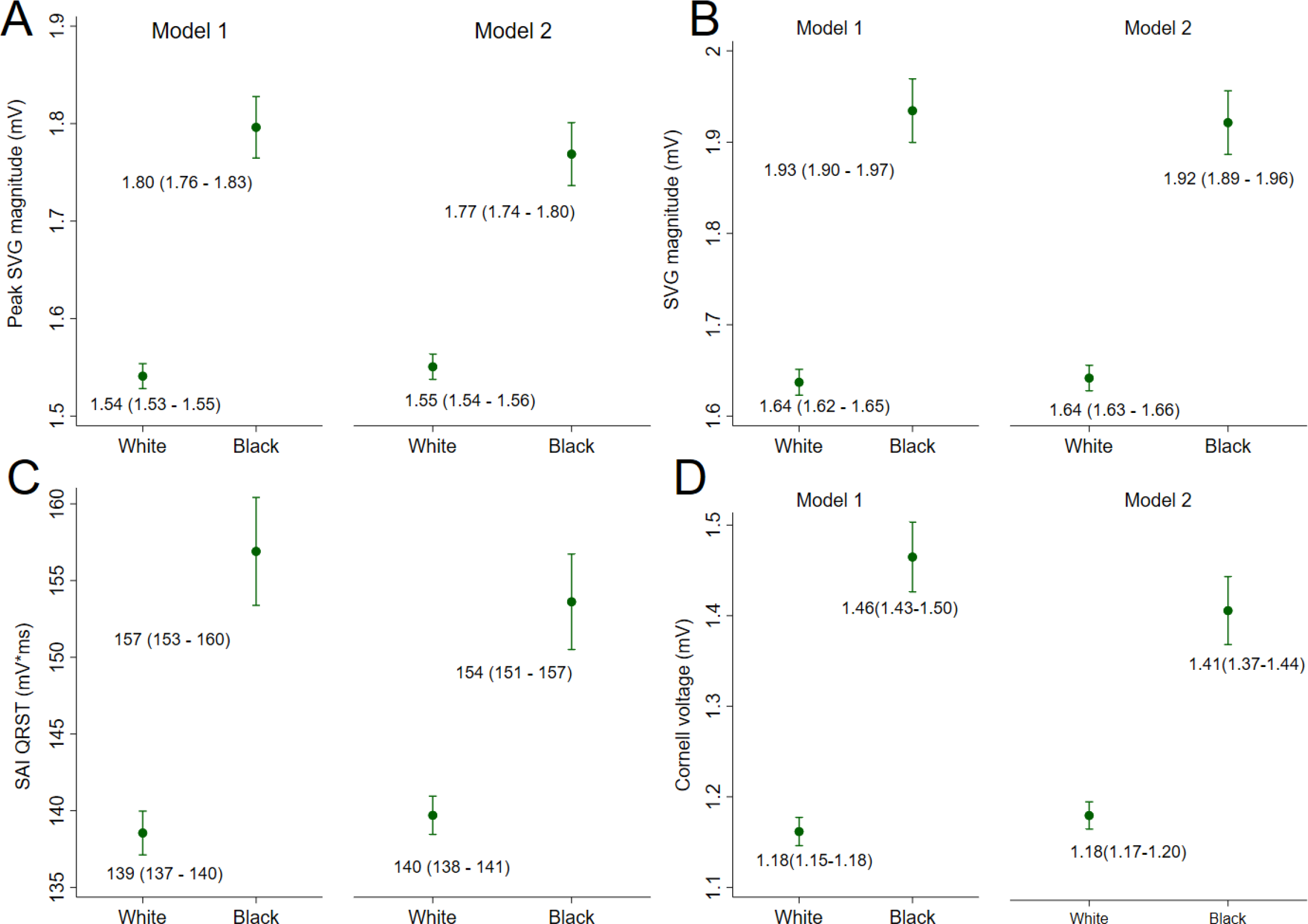
Estimated adjusted marginal (least-squares) means and 95% Confidence Intervals (CI) of (**A**) peak SVG magnitude, (**B**) area SVG magnitude, (**C**) SAI QRST, and (**D**) Cornell voltage for white and black participants. Model 1 was adjusted for age, sex, and study center. Model 2 was additionally adjusted for HF, CHD, stroke, diabetes, hypertension, current smoking and alcohol intake, work, sport, and leisure physical activity levels, levels of total cholesterol, high density lipoprotein, and triglycerides, BMI, use of antihypertensive and antiarrhythmic medications, serum concentrations of sodium, potassium, calcium, magnesium, phosphorus, and uric acid, total protein and albumin, blood urea nitrogen, and CKD stage classified by eGFR_CKD-EPI_, heart rate, QRS duration, Bazett-corrected QT interval, Cornell voltage, education level, occupation category, income, and health insurance.

### Association of EP substrate with sudden cardiac death in black and white participants

Over a median follow-up of 24.4 years, there were 522 SCDs (incidence 1.74; 95% CI 1.59-1.89 per 1000 person-years) and 2,147 nonSCDs (incidence 7.14; 95% CI 6.85-7.45 per 1000 person-years). SCD incidence was two times higher in black (2.86; 95%CI 2.50-3.28 per 1000 person-years) than white participants (1.37; 95%CI 1.22-1.53 per 1000 person-years). Incidence of nonSCD was higher in black (9.60; 95%CI 8.92-10.33 per 1000 person-years) than white participants (6.33; 95%CI 6.01-6.67 per 1000 person-years).

All traditional ECG metrics, SVG direction, and SAI QRST were associated with a similar risk of SCD in black and white individuals, a finding that was consistent both in Cox regression (Supplemental Table 1A) and competing risk models (Supplemental Table 2A).

Time-updated Cox regression model 2 revealed significant interaction with SVG magnitude. However, competing risk analysis found no effect modification by race of SVG magnitude – SCD association (Figure 2 and Supplemental Table 2). Continuous hazard functions looked similarly for white and black individuals (Supplemental Figure 2), suggesting an absence of meaningful effect modification.

**Figure 2.**
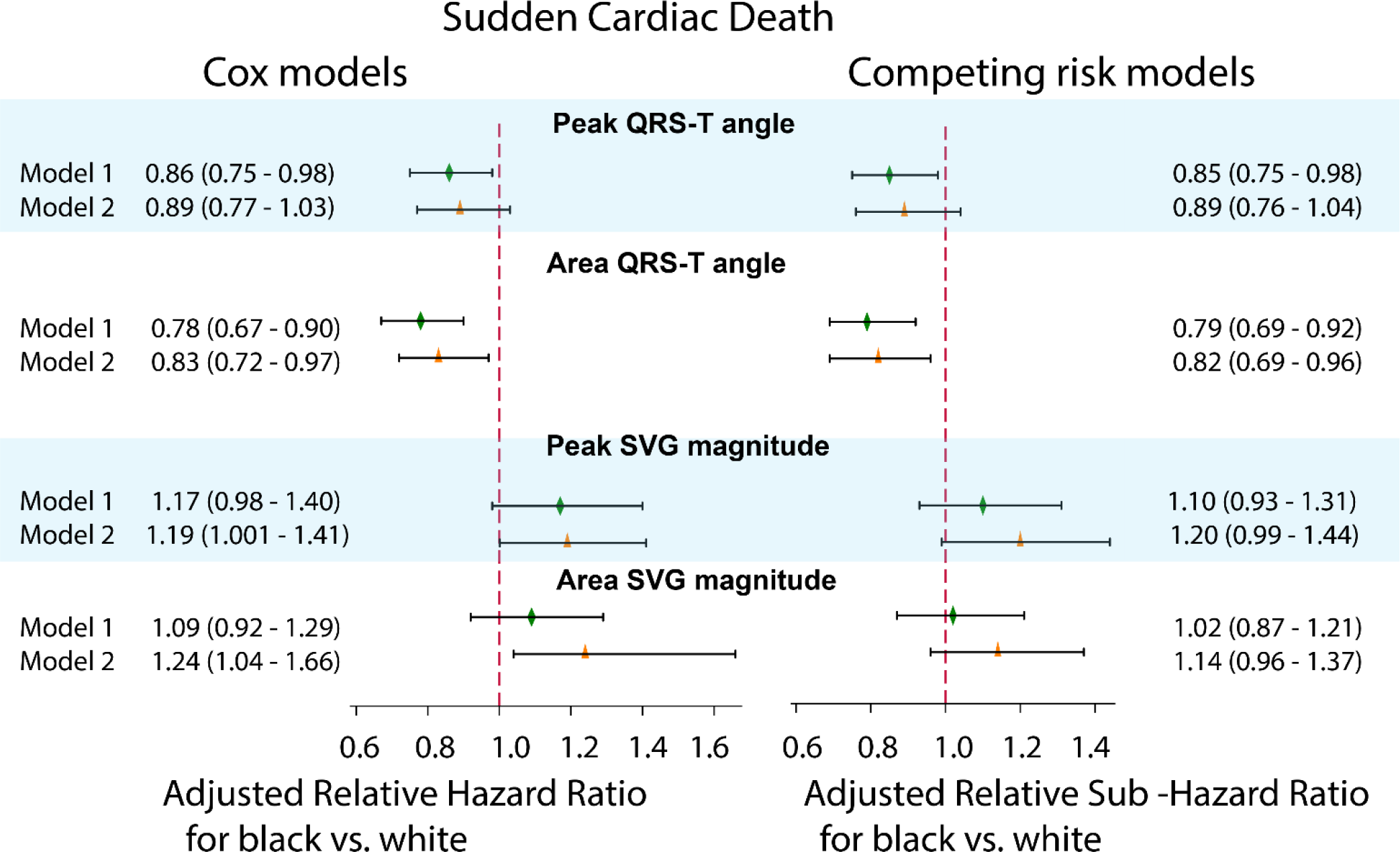
Adjusted Cox proportional relative hazard ratio (RHR) and competing SCD risk relative sub-hazard risk ratio (RSHR) with 95% confidence interval (CI) for black as compared to white participants, with HR/SHR for white participants equal 1.0. Models 1 (green diamond) and models 2 (orange triangle) for the QRS-T angle and SVG magnitude are shown. Black lines correspond to 95% CI bounds.

Race did not modify associations of ECG measures with nonSCD (Supplemental Table 2).

### Race and hypertension modify an association of spatial QRS-T angle with SCD

The lowest SCD incidence was observed in hypertension-free white (0.95; 95%CI 0.82-1.11 per 1000 person-years), followed by hypertension-free black (1.31; 95%CI 0.98-1.75 per 1000 person-years), and then by white individuals with hypertension (2.59; 95%CI 2.20-3.04 per 1000 person-years). Black individuals with hypertension had the highest rate of SCD: 4.26; 95%CI 3.66-4.96 per 1000 person-years.

Race significantly modified an association of spatial QRS-T angle with SCD, both in Cox regression (Figure 2 and Supplemental Table 1A) and competing risk models (Figure 2 and Supplemental Table 2A). In white individuals, the spatial QRS-T angle was associated with approximately 20% greater risk of SCD than in black individuals. Adjustment for incident nonfatal CVD in Cox model 2 attenuated the effect modification (Figure 2). In race-stratified Cox regression (Figure 3 and Supplemental Table 1B) and competing risk models 1 and 2 (Figure 4 and Supplemental Table 2B), spatial QRS-T angle was associated with SCD in white, but not black individuals (Figure 5).

**Figure 3.**
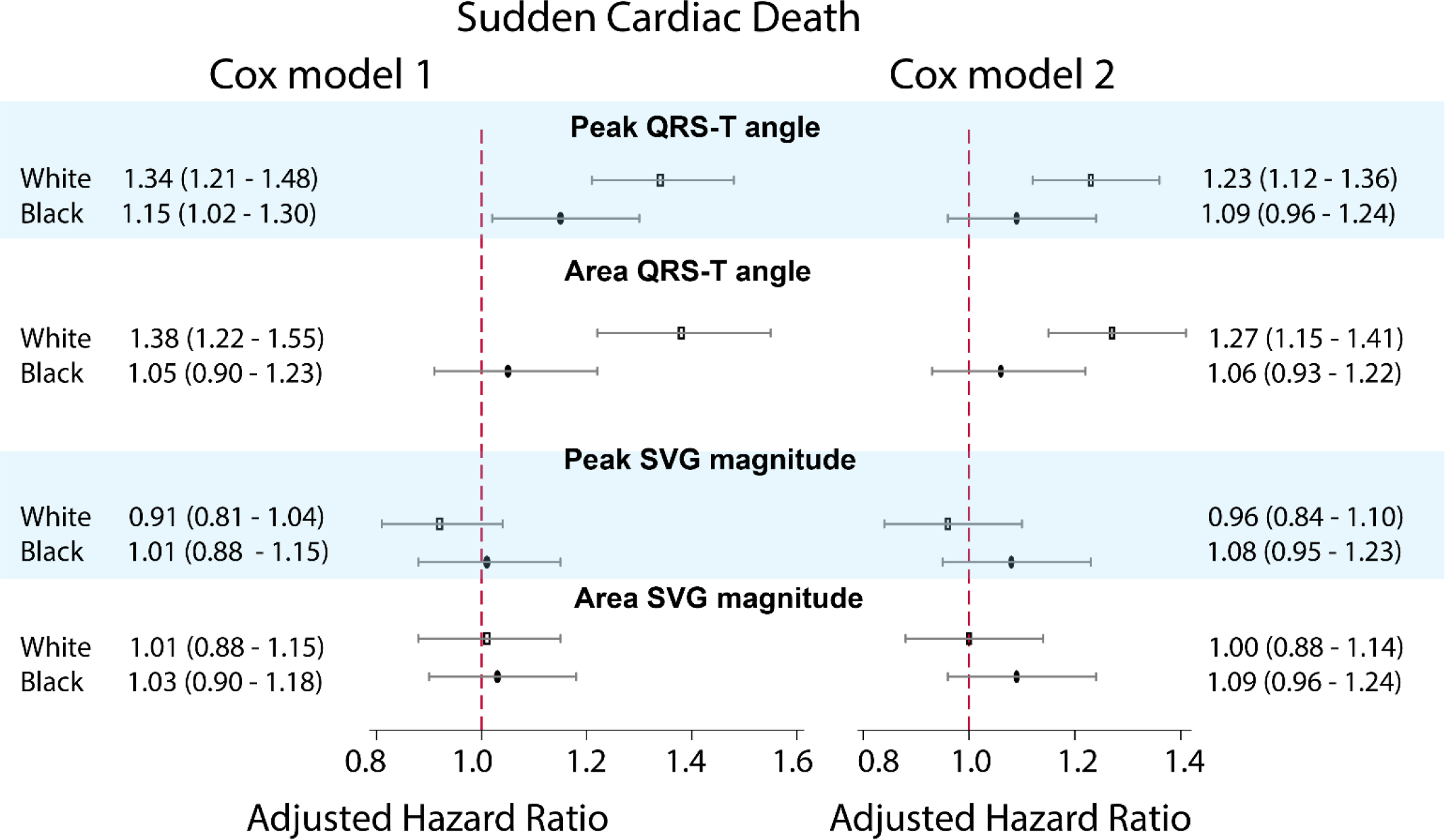
Sex-stratified adjusted (models 1 and 2) Cox proportional hazard ratio (HR) and 95% CI of SCD for QRS-T angle and SVG magnitude in white (hollow rectangle) and black (black oval) individuals. Black lines correspond to 95% CI bounds.

**Figure 4.**
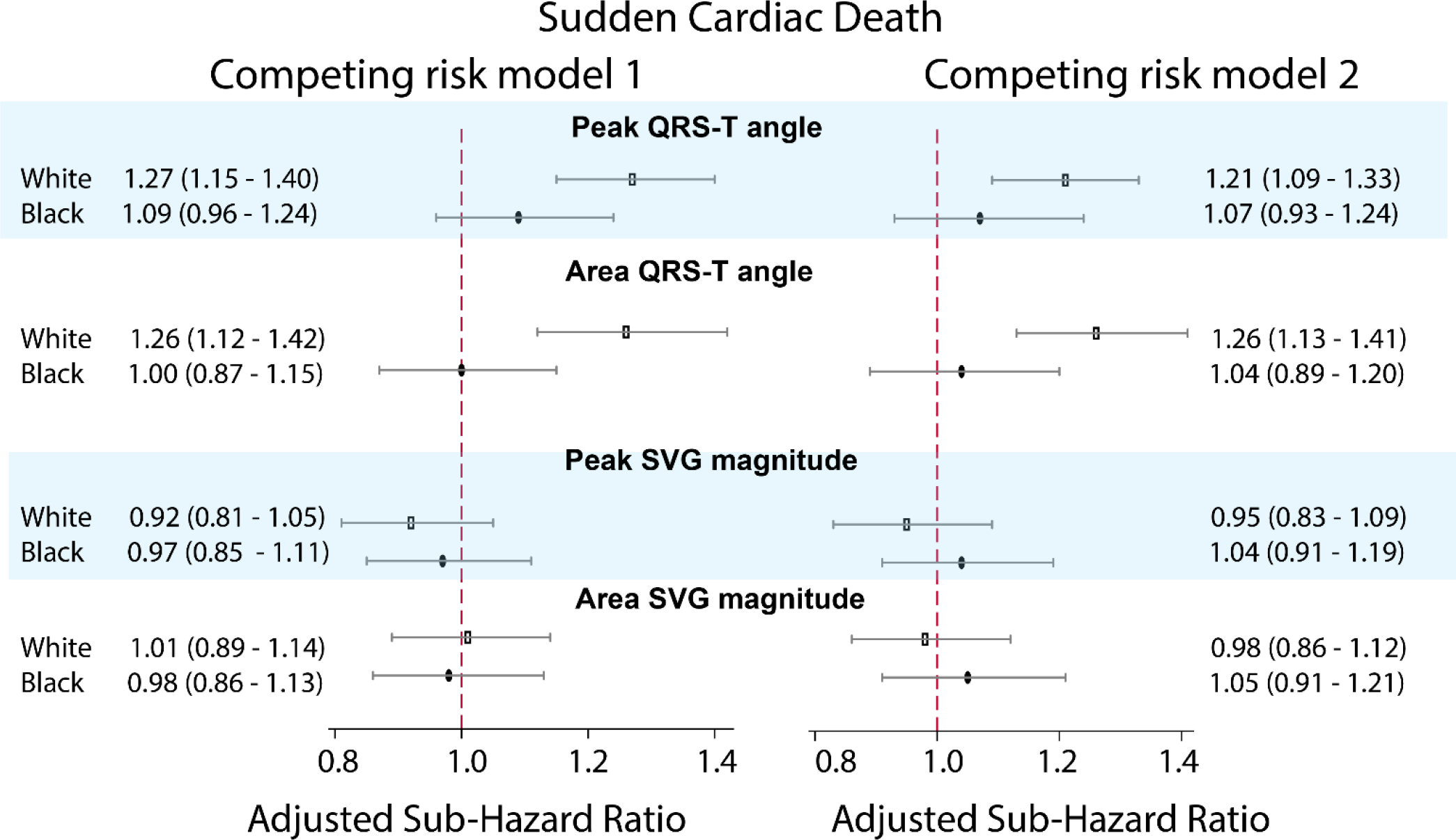
Adjusted (models 1 and 2) competing risk sub-hazard ratio (SHR) and 95% CI of SCD for QRS-T angle and SVG magnitude in white (hollow rectangle) and black (black oval) individuals. Black lines correspond to 95% CI bounds.

**Figure 5.**
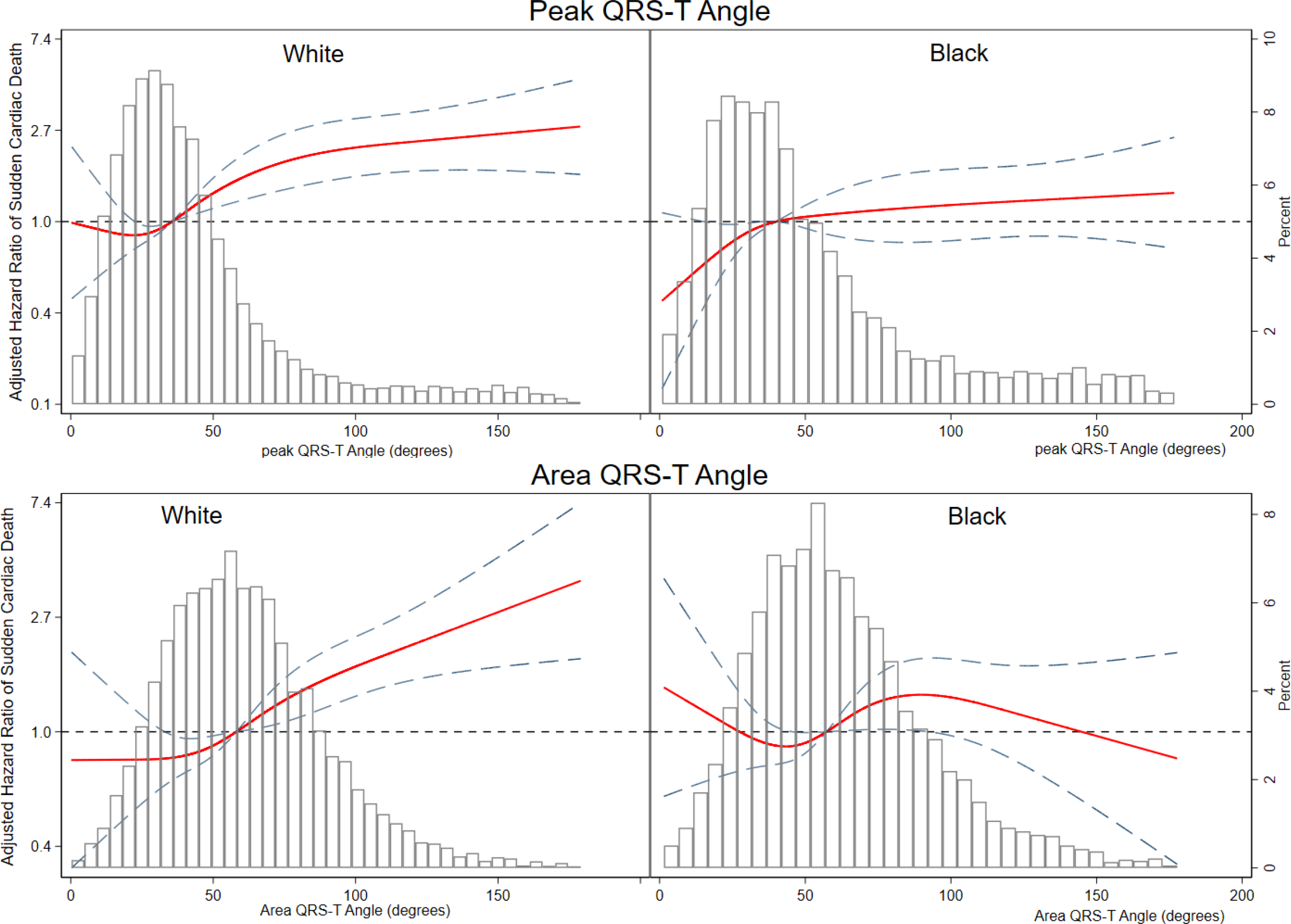
Adjusted (model 1) risk of SCD associated with area and peak QRS-T angle in black and white participants. Restricted cubic spline with 95% CI shows a change in the hazard ratio (Y-axis) in response to QRS-T angle change (X-axis). 50th percentile of QRS-T angle is selected as reference. Knots of peak QRS-T angle in white participants are at 10 – 28 – 44 -118 degrees, and in black participants are at 11 – 31 – 53 – 149 degrees. Knots of area QRS-T angle in white participants are at 22 – 48 – 69 – 112 degrees, and in black participants are at 21 – 48 – 69 - 118 degrees.

Further analysis of the interaction with both race and hypertension showed that spatial QRS-T angle was associated with SCD similarly in white and hypertension-free black individuals (Figure 6 and Supplemental Table 3), but not in black individuals with hypertension. Effect modification by race and hypertension remained significant after adjustment for incident CV events in both Cox regression and competing risk models.

**Figure 6.**
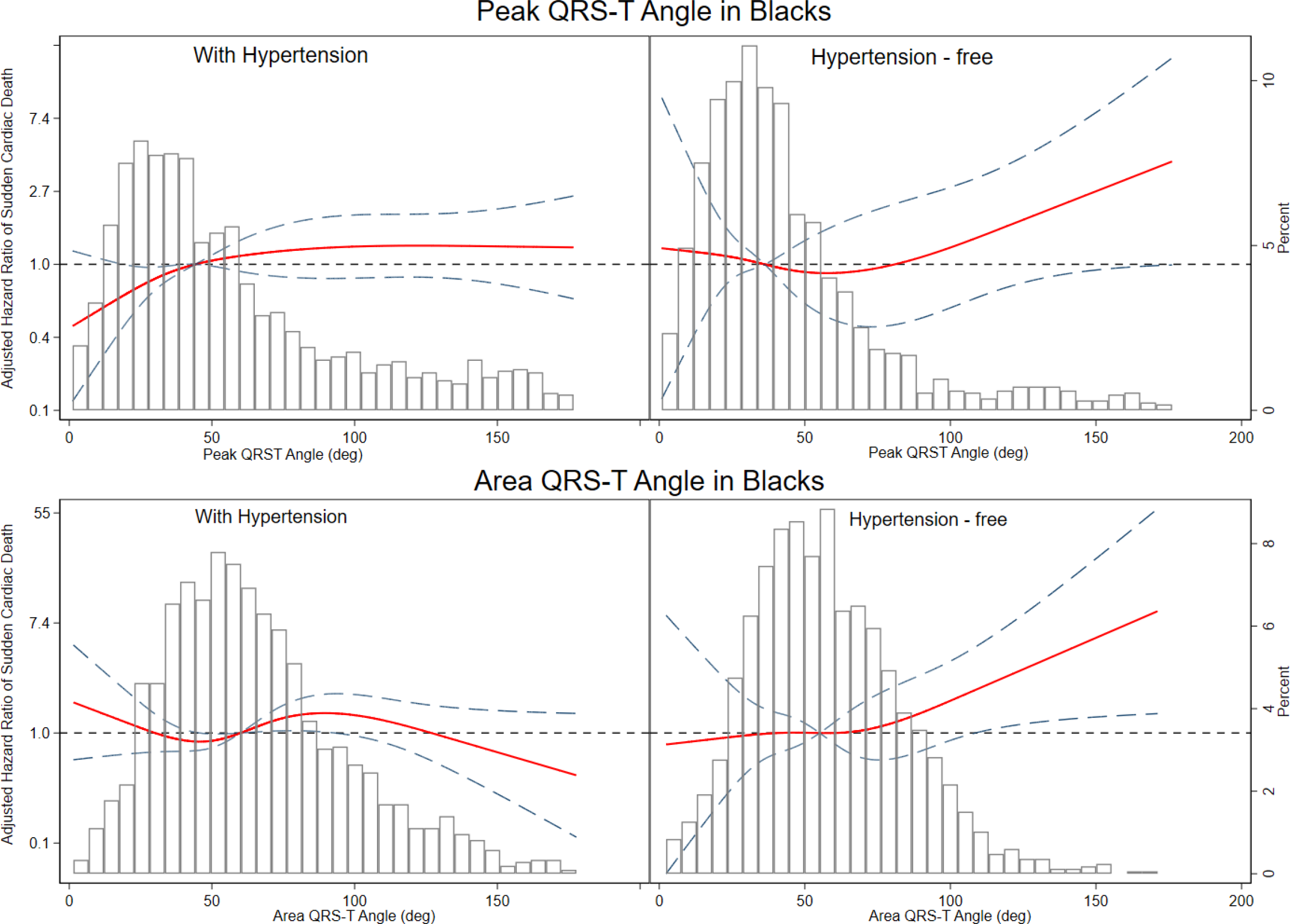
Adjusted (model 1) risk of SCD associated with area and peak QRS-T angle in black participants with and without hypertension. Restricted cubic spline with 95% CI shows a change in hazard ratio (Y-axis) in response to QRS-T angle change (X-axis). 50th percentile of QRS-T angle is selected as reference. Knots of peak QRS-T angle are at 10 – 29 – 45 - 120 degrees. Knots of area QRS-T angle in black participants with hypertension are at 21 – 50– 72 - 130 degrees, and in hypertension – free black participants are at 20 – 46 – 66 - 103 degrees.

## Discussion

This large, prospective community-based cohort study of more than 14,000 participants showed several important findings. Apart from the QRS-T angle, the association of EP substrate with SCD was similar in black and white individuals. In this study, race did not modify an association of EP substrate with SCD, supporting the recognition of race as a product of social practices, but not an inherent characteristic of individuals. Similar strength of the association of global ECG metrics with SCD in black and white individuals implies that EP substrate does not explain racial disparities in SCD rate.

Furthermore, we observed that black individuals with hypertension experienced 4.5-fold higher SCD incidence than hypertension-free white individuals. Black race in the presence of hypertension is associated with very high SCD risk regardless of QRS-T angle, which explains the weaker association of QRS-T angle with SCD in black than white individuals. However, relative risk of SCD carried by QRS-T angle did not differ in hypertension-free black and white individuals with or without hypertension. We also observed that black individuals had larger amplitude-based ECG metrics (SAI QRST, SVG magnitude, Cornell voltage) as compared to white individuals. The difference of approximately 0.3 mV was robustly observed even after accounting for socioeconomic status and other confounders. This finding is consistent with previous studies^35, 36^ and supports use of race-specific thresholds for abnormal SAI QRST,^8^ SVG magnitude,^8^ and ECG-LVH.^14, 37^

### Race does not modify an association of EP substrate with SCD

Our study found that all traditional ECG metrics (QTc duration, QRS duration, and Cornell voltage), SVG direction, and SAI QRST were associated with a similar risk of SCD among black and white individuals. There is little existing literature studying the association of ECG metrics and risk of SCD by race, and this study provides new insight into racial differences and similarities. In a prior study on sex differences,^32^ we found that Cornell voltage, SVG magnitude, and SAI QRST are associated with a 16-24% greater risk of SCD in women as compared to men. Sex is biologically determined and therefore may cause differences in EP substrate in contrast to race, which is a complex social construct.

There has been growing recognition of the Critical Race Theory^38^, which aims to better account for the effects of institutionalized racism on health. Critical Race Theory aims to acknowledge inequities in the biomedical field and bring to light both discrimination and health consequences that result from discrimination and inequalities in access to healthcare. Race is a product of social processes of power. The Critical Race Theory defines race not as an inherent characteristic of a person, but as a product of social practices.^38^ Results of our study support the Critical Race Theory postulate. Many previous studies recognized race as an independent and strong risk factor of SCD even after adjusting for socioeconomic factors but attributed it - at least partially – to underlying biological differences between black and white individuals.^1, 2, 4^ In contrast, our comprehensive study of EP substrate did not find meaningful effect modification by race. Results of our study call attention to structural racism as an important determinant of the increased SCD rate in black as compared to white individuals.^39^

A number of studies investigated the effects of racism on physical health. These studies showed that self-reported daily discrimination and stress was associated with increased waist circumference, increased waist to hip ratio, and higher fasting glucose level.^40-42^ Not only does daily stress increase potential risk factors for SCD, but long-term negative emotions also increase vulnerability to life-threatening arrhythmias^43^ and can potentially contribute to racial disparities in SCD.

When thinking about race and potential biological differences determining health, it is important to consider that race is not equivalent to genetic ancestry, but as mentioned above is a social construct. In our study, participants self-identified as black or white, but no ancestry information was analyzed. Ancestry analyses of the self-reported racial and ethnic identity in the US have shown that self-identified black individuals carry up to 24% of European ancestry, and about 1 in 10 self-identified white individuals in the US South have at least 1% of African ancestry.^44^ Association of genetic ancestry with EP substrate of SCD^4^ deserves separate investigation.

Clinical implications for the finding of no meaningful effect modification by race suggests that there is no need for race-specific risk stratification of SCD. Instead, race should be included in the SCD risk models as a potent risk marker.^8^

### Race-specific thresholds of ECG voltage measurements

Racial differences in ECG voltage and ECG-LVH definition have been previously reported.^35-37^ This study showed consistent results: voltage-based ECG metrics (SVG magnitude, SAI QRST, Cornell voltage) were larger in black than white individuals, by 0.2-0.3 mV.

Comprehensive adjustment for cardiovascular risk factors in this study did not affect the racial differences in SVG magnitude, SAI QRST, and Cornell voltage. Similarly, the Dallas Heart Study reported^45^ that after adjustment for cardiovascular risk factors and body composition, both black race and African ancestry were associated with approximately 0.25 mV larger ECG voltage. Of note, in our study, racial differences in SVG magnitude, SAI QRST, and Cornell voltage did not translate into different strengths of their associations with SCD.

A recent genome-wide association study of GEH revealed 10 GEH-associated loci.^46^ Four loci (11p11.2 cluster, near *ACTB, LUZP1-KDM1A*, and *IGF1R*) were associated with increase in both SAI QRST and SVG magnitude. *IGF1R* is a key signaling step in *physiologic* cardiomyocyte hypertrophy^47^, which can potentially explain why increased SAI QRST, SVG magnitude, and Cornell voltage in black individuals did not translate into stronger association of these ECG measures with SCD. SAI QRST – associated locus near *LUZP1-KDM1A* has higher effect allele frequency^46^ in African ancestry (0.92) as compared to European ancestry (0.59), which can potentially contribute to larger SAI QRST in a black population. Further studies are needed to test these hypotheses.

### Strengths and Limitations

The strengths of our study included the sizeable community-dwelling cohort, the extended follow-up, and the rigorous adjudication of SCD. We were also able to adjust for time-varying ECG measurements and incident nonfatal cardiovascular events and conduct competing risk analyses. However, our study has limitations, as previously acknowledged.^8^ While efforts were made to differentiate SCD from nonSCD, it is possible than some SCD events were secondary to sudden non-cardiac catastrophy as opposed to ventricular arrhythmias. However, these events likely compromise a small fraction of SCD events and therefore should not significantly affect study results. This study did not include resuscitated out-of-hospital cardiac arrest due to difficulty accurately differentiating its cause. Another important consideration is the inclusion of only black and white individuals, which limits generalizability to the multiethnic populations.

Further studies in other races and ethnicities are needed to validate our findings.

## Data Availability

The ARIC Study data are available through the National Heart, Lung, and Blood Institute’s Biological Specimen and Data Repository Information Coordinating Center (BioLINCC) and the National Center of Biotechnology Information’s database of Genotypes and Phenotypes (dbGaP). The MATLAB (MathWorks, Natick, MA, USA) open-source software code for GEH measurement and the heart vector origin definition is provided at https://physionet.org/physiotools/geh and https://github.com/Tereshchenkolab/Origin.

https://github.com/Tereshchenkolab/Origin

## Acknowledgments

The authors thank the staff and participants of the ARIC study for their important contributions. We would like to acknowledge the SCD mortality classification committee members: Nona Sotoodehnia (lead), Selcuk Adabag, Sunil Agarwal, Lin Chen, Rajat Deo, Leonard Ilkhanoff, Liviu Klein, Saman Nazarian, Ashleigh Owen, Kris Patton, and Larisa Tereshchenko.

## Funding Sources

The Atherosclerosis Risk in Communities study has been funded in whole or in part with Federal funds from the National Heart, Lung, and Blood Institute, National Institutes of Health, Department of Health and Human Services, under Contract nos. (HHSN268201700001I, HHSN268201700002I, HHSN268201700003I, HHSN268201700004I, HHSN268201700005I). This work was supported by HL118277 (LGT), and OHSU President Bridge funding (LGT).

## Disclosures

None.

---

**Supplemental Table 1A:** Race interaction in association of global ECG measures with SCD in Cox models

| Predictor, per 1 SD |  | All (n=14,408; 522 SCDs) |  |  |  |
| --- | --- | --- | --- | --- | --- |
|  |  | HR(95%CI) | P-value | RHR for black vs. white pts (95%CI) | P<sub>interaction</sub> |
| Model 1 | Peak QRS-T angle | **1.34(1.22-1.47)** | **<0.0001** | **0.86(0.75-0.98)** | **0.026** |
|  | Area QRS-T angle | **1.35(1.21-1.51)** | **<0.0001** | **0.78(0.67-0.90)** | **0.001** |
|  | Peak SVG elevation | **1.13(1.01-1.26)** | **0.027** | 0.92(0.78-1.09) | 0.348 |
|  | Area SVG elevation | **1.16(1.06-1.28)** | **0.002** | 0.88(0.75-1.04) | 0.136 |
|  | Peak SVG azimuth | **1.11(1.01-1.21)** | **0.026** | 1.01(0.88-1.15) | 0.940 |
|  | Area SVG azimuth | 0.99(0.90-1.09) | 0.878 | 1.07(0.92-1.25) | 0.389 |
|  | Peak SVG magnitude | 0.89(0.79-1.01) | 0.083 | 1.17(0.98-1.40) | 0.083 |
|  | Area SVG magnitude | 0.98(0.86-1.11) | 0.753 | 1.09(0.92-1.29) | 0.314 |
|  | SAI QRST | **1.21(1.08-1.35)** | **0.001** | 0.89(0.79-1.01) | 0.078 |
|  | Heart rate | 1.11(0.99-1.24) | 0.081 | 1.01(0.86-1.19) | 0.907 |
|  | Bazett's QTc | **1.12(1.01-1.24)** | **0.032** | 1.01(0.89-1.14) | 0.924 |
|  | QRS duration | 1.07(0.98-1.18) | 0.128 | 0.93(0.81-1.07) | 0.304 |
|  | Cornell voltage | 1.10(0.98-1.24) | 0.104 | 1.01(0.87-1.18) | 0.893 |
| Model 2 | Peak QRS-T angle | **1.24(1.12-1.36)** | **<0.0001** | 0.89(0.77-1.03) | 0.121 |
|  | Area QRS-T angle | **1.27(1.15-1.40)** | **<0.0001** | **0.83(0.72-0.97)** | **0.020** |
|  | Peak SVG elevation | **1.14(1.03-1.27)** | **0.011** | 0.96(0.81-1.15) | 0.675 |
|  | Area SVG elevation | **1.12(1.01-1.23)** | **0.035** | 0.97(0.81-1.16) | 0.726 |
|  | Peak SVG azimuth | **1.12(1.02-1.22)#** | **0.015** | 1.04(0.90-1.21) | 0.602 |
|  | Area SVG azimuth | 1.04(0.95-1.15) | 0.387 | 1.11(0.93-1.32) | 0.246 |
|  | Peak SVG magnitude | 0.98(0.86-1.11) | 0.720 | **1.19(1.001-1.41)** | **0.048** |
|  | Area SVG magnitude | 0.93(0.82-1.07) | 0.310 | **1.24(1.04-1.66)** | **0.018** |
|  | SAI QRST | **1.10(1.02-1.19)** | **0.013** | 1.01(0.91-1.12) | 0.865 |
|  | Heart rate | 1.11(0.99-1.24) | 0.078 | 1.04(0.89-1.23) | 0.600 |
|  | Bazett's QTc | **1.16(1.06-1.27)#** | **0.002** | 1.01(0.88-1.15) | 0.916 |
|  | QRS duration | **1.18(1.09-1.29)** | **<0.0001** | 0.96(0.84-1.10) | 0.575 |
|  | Cornell voltage | 1.06(0.97-1.16) | 0.187 | 1.09(0.97-1.23) | 0.135 |

#Proportionality hazards assumption not met; SVG=spatial ventricular gradient: RHR=relative hazard ratio.

**Supplemental Table 1B:** Association of GEH with SCD in Cox models for white and black participants

| Predictor, per 1 SD |  | White (n=10,669; 309 SCDs) |  | Black (n=3,739; 213 SCDs) |  |
| --- | --- | --- | --- | --- | --- |
|  |  | HR(95% CI) | P-value | HR(95% CI) | P-value |
| Model 1 | Peak QRS-T angle | **1.34(1.21-1.48)** | **<0.0001** | **1.15(1.02-1.30)** | **0.018** |
|  | Area QRS-T angle | **1.38(1.22-1.55)** | **<0.0001** | 1.05(0.91-1.22) | 0.467 |
|  | Peak SVG elevation | **1.13(1.01-1.27)** | **0.032** | 1.05(0.90-1.23) | 0.523 |
|  | Area SVG elevation | **1.17(1.06-1.30)** | **0.002** | 1.05(0.90-1.21) | 0.555 |
|  | Peak SVG azimuth | **1.10(1.003-1.21)** | **0.044** | 1.10(0.97-1.26) | 0.129 |
|  | Area SVG azimuth | 0.99(0.90-1.10) | 0.901 | 1.05(0.91-1.22) | 0.471 |
|  | Peak SVG magnitude | 0.92(0.81-1.04) | 0.198 | 1.01(0.88-1.15) | 0.915 |
|  | Area SVG magnitude | 1.01(0.88-1.15) | 0.935 | 1.03(0.90-1.18) | 0.625 |
|  | SAI QRST | **1.23(1.08-1.39)** | **0.001** | 1.07(0.95-1.21) | 0.270 |
|  | Heart rate | 1.11(0.98-1.26) | 0.090 | **1.16(1.02-1.32)** | **0.029** |
|  | Bazett's QTc | **1.18(1.03-1.29)** | **0.010** | **1.11(1.002-1.23)** | **0.046** |
|  | QRS duration | 1.06(0.95-1.18) | 0.272 | 1.04(0.92-1.18) | 0.510 |
|  | Cornell voltage | 1.07(0.95-1.22) | 0.256 | **1.15(1.02-1.30)** | **0.018** |
| Model 2 | Peak QRS-T angle | **1.23(1.12-1.36)** | **<0.0001** | 1.09(0.96-1.24) | 0.171 |
|  | Area QRS-T angle | **1.27(1.15-1.41)** | **<0.0001** | 1.06(0.93-1.22) | 0.373 |
|  | Peak SVG elevation | **1.16(1.04-1.29)** | **0.006** | 1.12(0.96-1.30) | 0.147 |
|  | Area SVG elevation | **1.13(1.02-1.25)** | **0.021** | 1.09(0.94-1.28) | 0.251 |
|  | Peak SVG azimuth | **1.11(1.01-1.21)** | **0.022** | **1.16(1.02-1.32)** | **0.023** |
|  | Area SVG azimuth | **1.05(0.95-1.16)** | **0.359** | 1.15(0.99-1.34) # | 0.074 |
|  | Peak SVG magnitude | 0.96(0.84-1.10) | 0.595 | 1.08(0.95-1.23) | 0.236 |
|  | Area SVG magnitude | 1.00(0.88-1.14) | 0.964 | 1.09(0.96-1.24) | 0.183 |
|  | SAI QRST | **1.13(1.04-1.22)** | **0.004** | **1.11(1.01-1.21)** | **0.023** |
|  | Heart rate | 1.09(0.97-1.23) | 0.134 | **1.20(1.06-1.36)** | **0.005** |
|  | Bazett's QTc | **1.19(1.09-1.30)** | **<0.0001** | **1.16(1.05-1.29)** | **0.004** |
|  | QRS duration | **1.18(1.08-1.29)** | **<0.0001** | **1.15(1.03-1.30)** | **0.015** |
|  | Cornell voltage | 1.08(0.99-1.19) | 0.082 | **1.16(1.06-1.27)** | **0.001** |

#Proportionality hazards assumption not met; SVG=spatial ventricular gradient.

**Supplemental Table 2A.** Race interaction in association of GEH with SCD and nonSCD in competing risk models

| Predictor, per 1 SD |  | SCD (n=14,408; 522 SCDs) |  |  |  | nonSCD (n=14,408; 2,147 nonSCDs) |  |  |  |
| --- | --- | --- | --- | --- | --- | --- | --- | --- | --- |
|  |  | SHR(95% CI) | P-value | RSHR for black vs.white (95% CI) | P<sub>interaction</sub> | SHR(95% CI) | P-value | RSHR for black vs.white (95% CI) | P<sub>interaction</sub> |
| Model 1 | Peak QRS-T angle | **1.28(1.16-1.40)** | **<0.0001** | **0.85(0.75-0.98)** | **0.022** | **1.11(1.05-1.17)** | **<0.0001** | 1.01(0.93-1.09) | 0.979 |
|  | Area QRS-T angle | **1.25(1.13-1.39)** | **<0.0001** | **0.79(0.69-0.92)** | **0.002** | **1.15(1.08-1.22)** | **<0.0001** | 1.00(0.91-1.09) | 0.914 |
|  | Peak SVG elevation | 1.11(0.99-1.25) | 0.075 | 0.96(0.80-1.14) | 0.630 | 1.00(0.95-1.06) | 0.869 | 0.930(0.84-1.03) | 0.148 |
|  | Area SVG elevation | **1.14(1.03-1.25)** | **0.010** | 0.94(0.80-1.11) | 0.471 | 1.00(0.94-1.05) | 0.904 | 0.96(0.87-1.05) | 0.398 |
|  | Peak SVG azimuth | 1.09(0.99-1.19) | 0.071 | 1.02(0.89-1.18) | 0.755 | 1.04(0.99-1.09) | 0.168 | 0.99(0.91-1.09) | 0.890 |
|  | Area SVG azimuth | 0.98(0.88-1.08) | 0.640 | 1.07(0.91-1.64) | 0.426 | **1.07(1.01-1.13)** | **0.017** | 0.99(0.90-1.09) | 0.837 |
|  | Peak SVG magnitude | 0.90(0.79-1.02) | 0.096 | 1.10(0.93-1.31) | 0.263 | 0.96(0.90-1.02) | 0.170 | 1.10(0.99-1.21) | 0.073 |
|  | Area SVG magnitude | 0.98(0.87-1.11) | 0.727 | 1.02(0.87-1.21) | 0.792 | 0.98(0.92-1.05) | 0.608 | 1.08(0.98-1.19) | 0.114 |
|  | SAI QRST | **1.13(1.02-1.26)** | **0.019** | 0.88(0.77-1.01) | 0.076 | 1.05(0.98-1.13) | 0.192 | 1.01(0.93-1.10) | 0.751 |
|  | Heart rate. | 1.03(0.92-1.16) | 0.576 | 0.99(0.84-1.17) | 0.918 | **1.13(1.07-1.20)** | **<0.0001** | 1.07(0.98-1.17) | 0.128 |
|  | Bazett's QTc | 1.09(0.98-1.21) | 0.124 | 1.02(0.90-1.17) | 0.737 | 1.05(0.996-1.11) | 0.071 | 1.02(0.94-1.09) | 0.673 |
|  | QRS duration | 1.06(0.95-1.17) | 0.293 | 0.92(0.78-1.07) | 0.267 | 1.05(0.996-1.11) | 0.069 | 0.98(0.90-1.06) | 0.608 |
|  | Cornell voltage | 1.08(0.96-1.22) | 0.185 | 0.98(0.84-1.15) | 0.815 | **1.12(1.05-1.20)** | **0.001** | 0.97(0.89-1.07) | 0.543 |
| Model 2 | Peak QRS-T angle | **1.21(1.10-1.33)** | **<0.0001** | 0.89(0.76-1.04) | 0.130 | **0.93(0.87-0.98)** | **0.010** | 1.02(0.93-1.12) | 0.706 |
|  | Area QRS-T angle | **1.26(1.13-1.41)** | **<0.0001** | **0.82(0.69-0.96)** | **0.017** | **0.92(0.87-0.97)** | **0.004** | 1.02(0.92-1.13) | 0.656 |
|  | Peak SVG elevation | **1.16(1.04-1.29)** | **0.007** | 0.96(0.80-1.15) | 0.649 | **0.92(0.86-0.98)** | **0.006** | 1.07(0.95-1.21) | 0.281 |
|  | Area SVG elevation | **1.11(1.005-1.24)** | **0.041** | 0.98(0.82-1.17) | 0.849 | **0.93(0.88-0.99)** | **0.031** | 1.05(0.93-1.18) | 0.418 |
|  | Peak SVG azimuth | **1.11(1.01-1.22)** | **0.033** | 1.04(0.88-1.23) | 0.651 | **0.95(0.90-0.999)** | **0.044** | 1.01(0.91-1.12) | 0.887 |
|  | Area SVG azimuth | 1.04(0.93-1.16) | 0.458 | 1.10(0.89-1.81) | 0.360 | 0.96(0.91-1.02) | 0.171 | 0.97(0.87-1.07) | 0.549 |
|  | Peak SVG magnitude | 0.91(0.79-1.04) | 0.180 | 1.20(0.99-1.44) | 0.058 | 1.04(0.97-1.12) | 0.273 | 0.94(0.84-1.05) | 0.248 |
|  | Area SVG magnitude | 0.99(0.83-1.08) | 0.425 | 1.14(0.96-1.37) | 0.144 | 1.04(0.97-1.12) | 0.273 | 0.93(0.83-1.03) | 0.179 |
|  | SAI QRST | **1.10(1.02-1.19)** | **0.011** | 0.99(0.87-1.12) | 0.831 | 0.95(0.90-1.01) | 0.083 | 0.97(0.89-1.05) | 0.447 |
|  | Heart rate. | 1.03(0.92-1.15) | 0.589 | 1.05(0.88-1.25) | 0.583 | **1.11(1.04-1.18)** | **0.001** | 0.97(0.88-1.08) | 0.590 |
|  | Bazett's QTc | **1.13(1.03-1.24)** | **0.009** | 1.02(0.89-1.16) | 0.792 | 1.02(0.97-1.09) | 0.413 | 0.96(0.88-1.06) | 0.418 |
|  | QRS duration | **1.20(1.10-1.30)** | **<0.0001** | 0.96(0.83-1.10) | 0.530 | **0.92(0.87-0.97)** | **0.002** | 0.98(0.89-1.08) | 0.713 |
|  | Cornell voltage | 1.05(0.96-1.15) | 0.257 | 1.09(0.96-1.24) | 0.204 | 0.97(0.92-1.02) | 0.187 | 1.00(0.92-1.08) | 0.937 |

RSHR=relative sub-hazard ratio

**Supplemental Table 2B.**
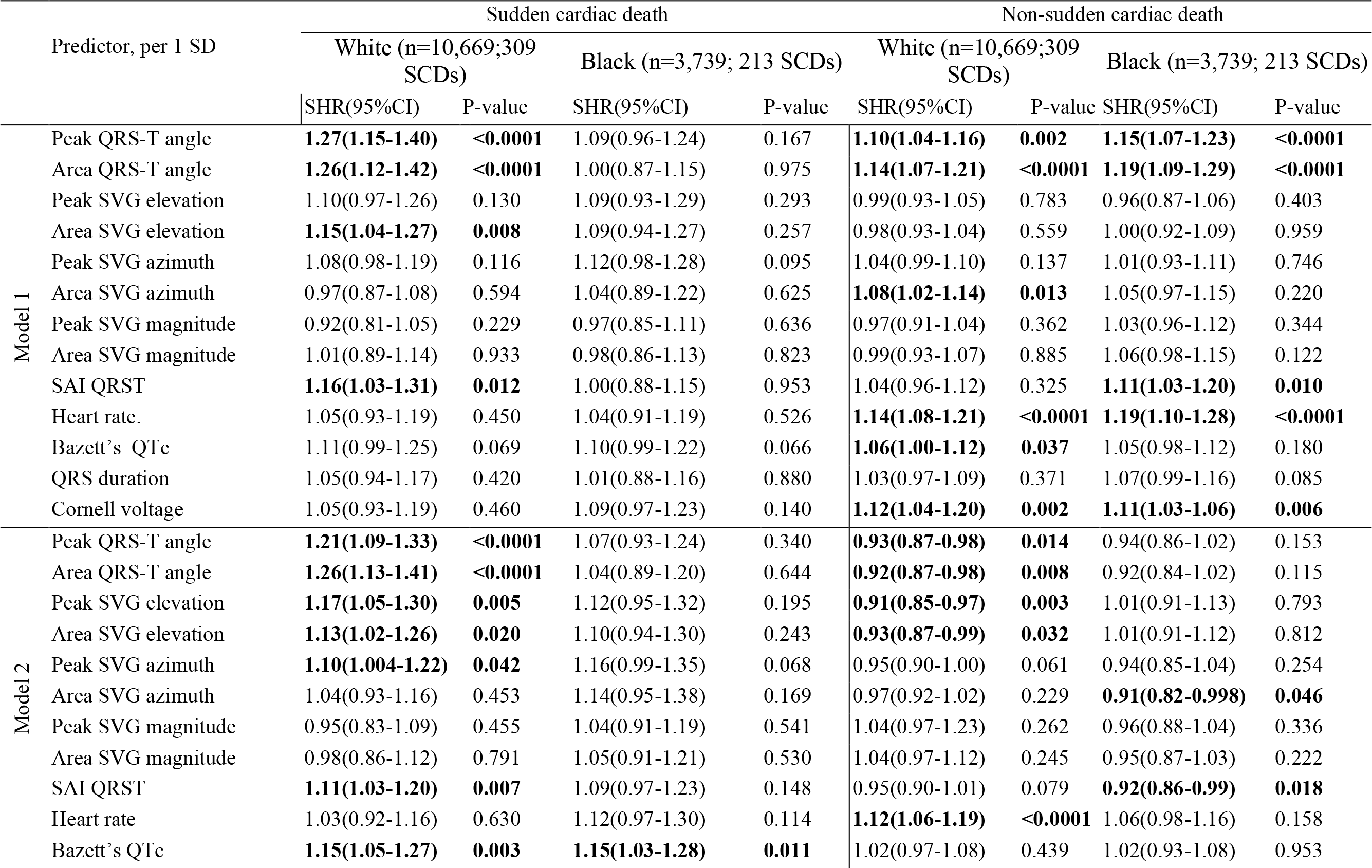
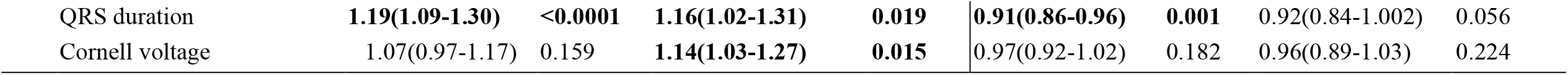
Competing risks of sudden cardiac death and non-sudden cardiovascular death for white and black participants

**Supplemental Table 3.** Relative risk of SCD associated with SVG magnitude and QRS-T angle in race-hypertension categories

|  | Race-Hypertension category | Peak QRS-T angle, per 1SD |  | Area QRS-T angle, per 1SD |  | Peak SVG magnitude, per 1SD |  | Area SVG magnitude, per 1SD |  |
| --- | --- | --- | --- | --- | --- | --- | --- | --- | --- |
|  |  | RHR(95%CI) | P-value | RHR(95%CI) | P-value | RHR(95%CI) | P-value | RHR(95%CI) | P-value |
| Cox Model 1 | White HTN-free (n=7,784) | Reference (1) |  | Reference (1) |  | Reference (1) |  | Reference (1) |  |
|  | White with HTN (n=2,885) | 0.94(0.79-1.12) | 0.507 | 0.93(0.77-1.13)# | 0.458 | 1.26(0.98-1.61)# | 0.066 | 1.28(0.99-1.64)# | 0.055 |
|  | Black with HTN (n=2,077) | **0.82(0.70-0.97)** | **0.021** | **0.71(0.59-0.86)** | **<0.0001** | **1.32(1.05-1.65)** | **0.016** | 1.25(0.999-1.56) | 0.051 |
|  | Black HTN-free (n=1,662) | 0.87(0.66-1.14) | 0.304 | 1.04(0.71-1.53) | 0.681 | **1.44(1.04-2.01)** | **0.030** | 1.35(0.98-1.85) | 0.070 |
| Cox Model 2 | White HTN-free (n=7,784) | Reference (1) |  | Reference (1) |  | Reference (1) |  | Reference (1) |  |
|  | White with HTN (n=2,885) | 0.88(0.74-1.06) | 0.170 | 0.92(0.76-1.10) | 0.359 | 1.18(0.91-1.54) | 0.203 | 1.17(0.91-1.51) | 0.218 |
|  | Black with HTN (n=2,077) | **0.82(0.69-0.98)** | **0.029** | **0.76(0.62-0.91)** | **0.004** | **1.30(1.03-1.63)** | **0.026** | 1.25(0.999-1.56) | 0.051 |
|  | Black HTN-free (n=1,662) | 0.83(0.62-1.12) | 0.229 | 0.93(0.68-1.26) | 0.631 | **1.49(1.08-2.05)** | **0.014** | **1.41(1.03-1.92)** | **0.031** |
|  |  | RSHR(95%CI) | P-value | RSHR(95%CI) | P-value | RSHR(95%CI) | P-value | RSHR(95%CI) | P-value |
| Competing SCD model 1 | White HTN-free (n=7,784) | Reference (1) |  | Reference (1) |  | Reference (1) |  | Reference (1) |  |
|  | White with HTN (n=2,885) | 0.93(0.78-1.09) | 0.368 | 0.93(0.77-1.12) | 0.448 | 1.26(0.99-1.60) | 0.063 | **1.27(1.01-1.61)** | **0.043** |
|  | Black with HTN (n=2,077) | **0.79(0.67-0.93)** | **0.005** | **0.71(0.59-0.86)** | **<0.0001** | 1.19(0.96-1.48) | 0.120 | 1.12(0.90-1.38) | 0.314 |
|  | Black HTN-free (n=1,662) | 0.93(0.71-1.23) | 0.611 | 1.04(0.74-1.44) | 0.834 | **1.42(1.07-1.89)** | **0.017** | **1.32(1.00-1.76)** | **0.050** |
| Competing SCD model 2 | White HTN-free (n=7,784) | Reference (1) |  | Reference (1) |  | Reference (1) |  | Reference (1) |  |
|  | White with HTN (n=2,885) | 0.87(0.72-1.04) | 0.134 | 0.90(0.74-1.10) | 0.320 | 1.15(0.87-1.51) | 0.326 | 1.16(0.90-1.51) | 0.255 |
|  | Black with HTN (n=2,077) | **0.80(0.67-0.96)** | **0.019** | **0.72(0.59-0.89)** | **0.002** | 1.22(0.97-1.53) | 0.090 | 1.18(0.94-1.48) | 0.155 |
|  | Black HTN-free (n=1,662) | 0.87(0.64-1.17) | 0.348 | 0.97(0.71-1.33) | 0.850 | **1.50(1.07-2.09)** | **0.018** | **1.44(1.05-1.96)** | **0.023** |
| Competing nonSCD model 1 | White HTN-free (n=7,784) | Reference (1) |  | Reference (1) |  | Reference (1) |  | Reference (1) |  |
|  | White with HTN (n=2,885) | 0.90(0.81-0.99) | 0.032 | 0.90(0.82-1.01) | 0.066 | 0.97(0.85-1.10) | 0.599 | 0.98(0.86-1.11) | 0.730 |
|  | Black with HTN (n=2,077) | 0.94(0.85-1.04) | 0.259 | 0.94(0.84-1.05) | 0.303 | 1.10(0.98-1.25) | 0.112 | 1.09(0.96-1.22) | 0.172 |
|  | Black HTN-free (n=1,662) | 0.98(0.65-1.13) | 0.764 | 0.93(0.79-1.11) | 0.435 | 0.97(0.80-1.16) | 0.708 | 0.99(0.83-1.18) | 0.890 |
| Competing nonSCD model 2 | White HTN-free (n=7,784) | Reference (1) |  | Reference (1) |  | Reference (1) |  | Reference (1) |  |
|  | White with HTN (n=2,885) | 0.93(0.83-1.04) | 0.219 | 0.93(0.83-1.03) | 0.164 | 0.95(0.83-1.10) | 0.493 | 0.88(0.76-1.00) | 0.056 |
|  | Black with HTN (n=2,077) | 0.96(0.86-1.09) | 0.558 | 0.99(0.87-1.12) | 0.814 | 0.92(0.81-1.05) | 0.235 | 0.88(0.76-1.00) | 0.053 |
|  | Black HTN-free (n=1,662) | 1.02(0.87-1.20) | 0.797 | 0.97(0.80-1.16) | 0.724 | 0.88(0.74-1.05) | 0.146 | **0.83(0.70-0.99)** | **0.042** |

## Supplemental Figure Legends

**Supplemental Figure 1:**
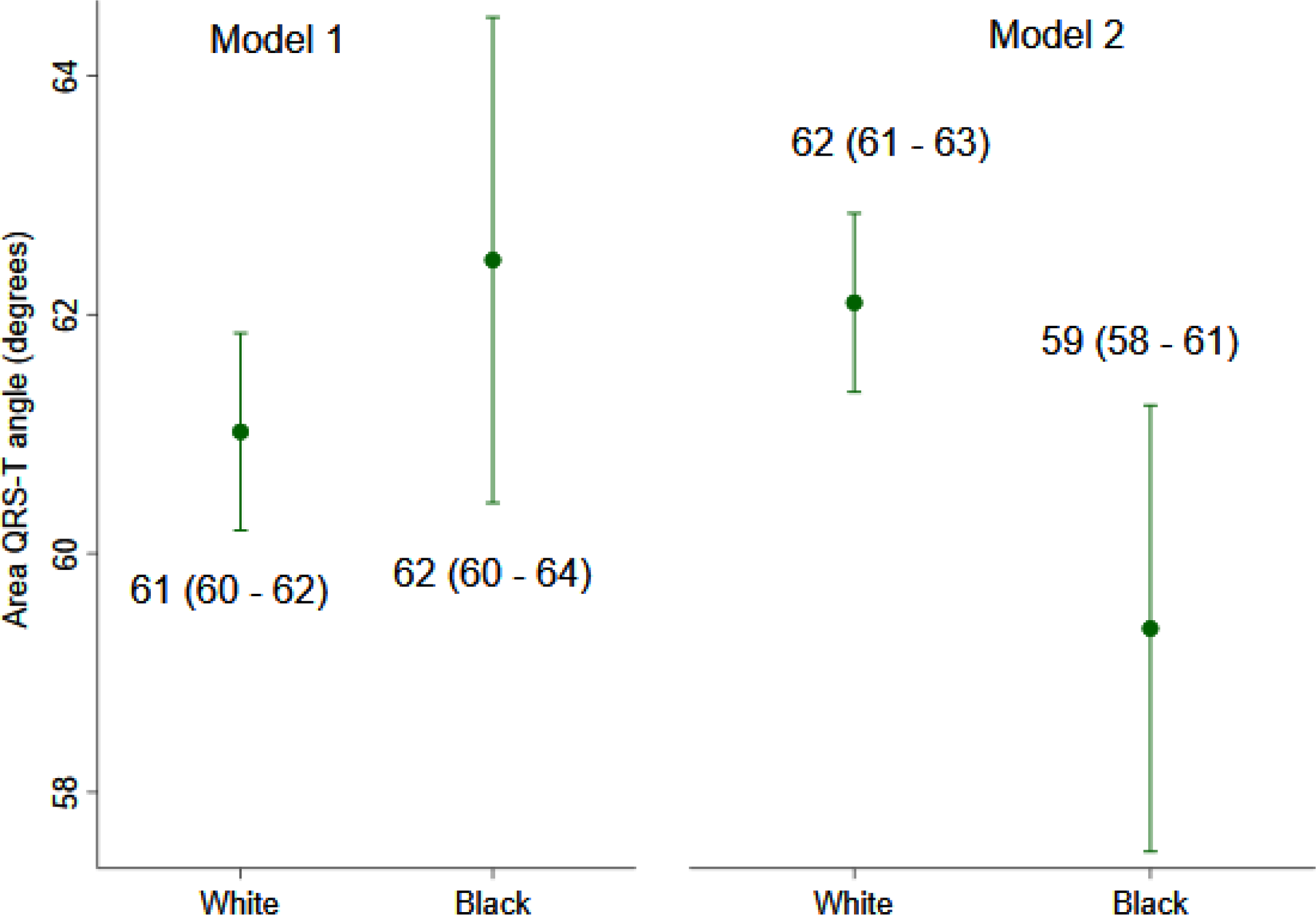
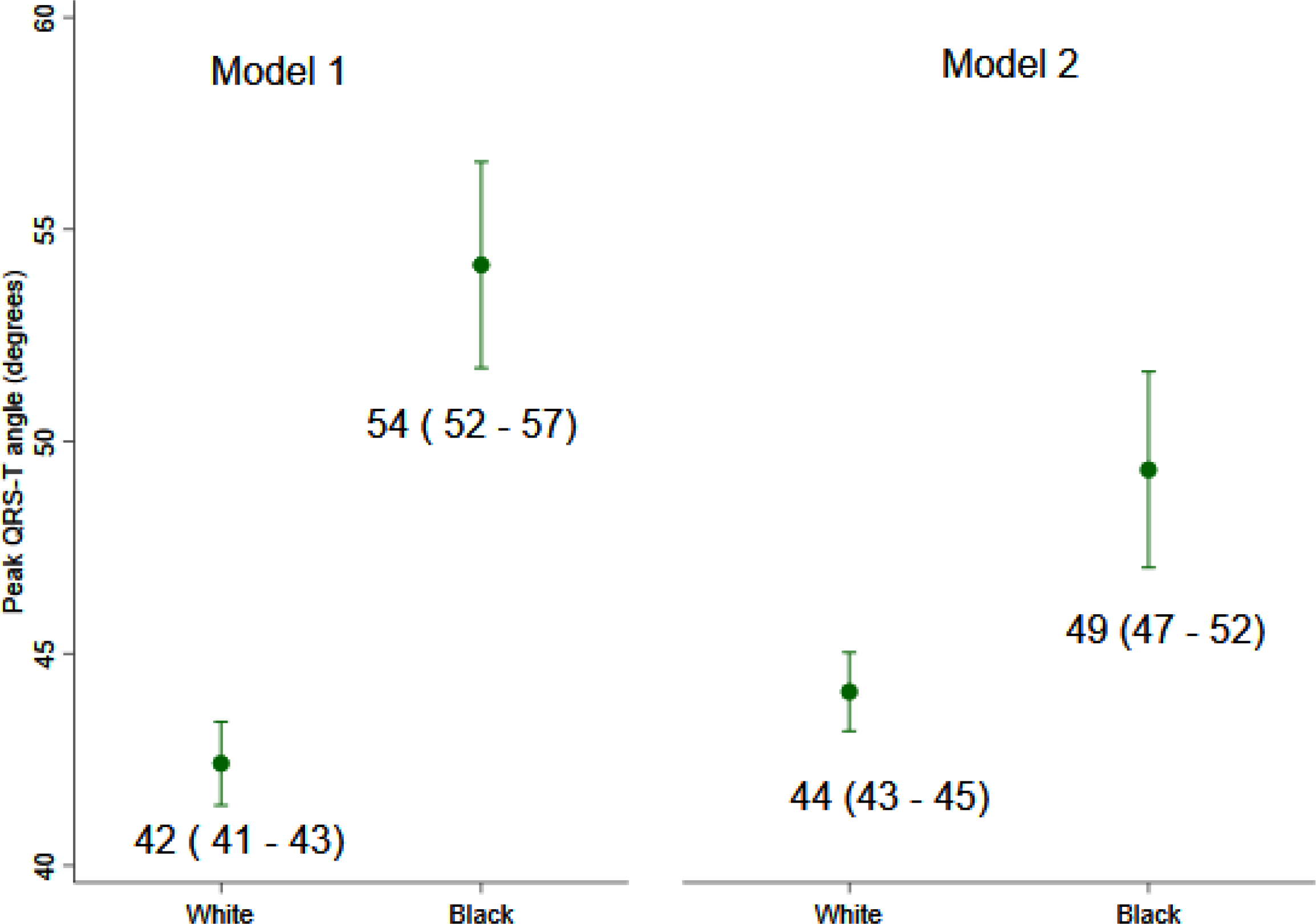
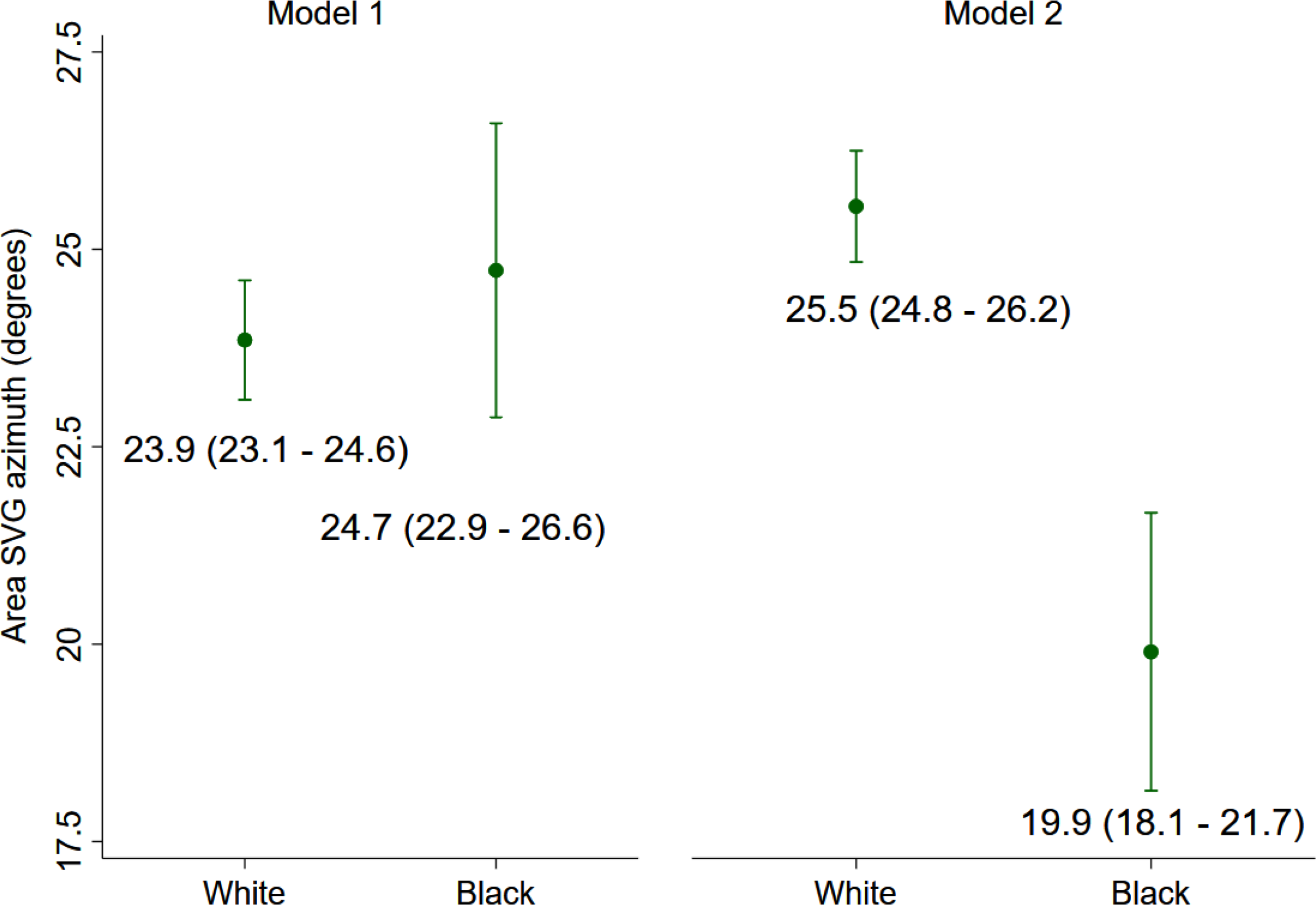
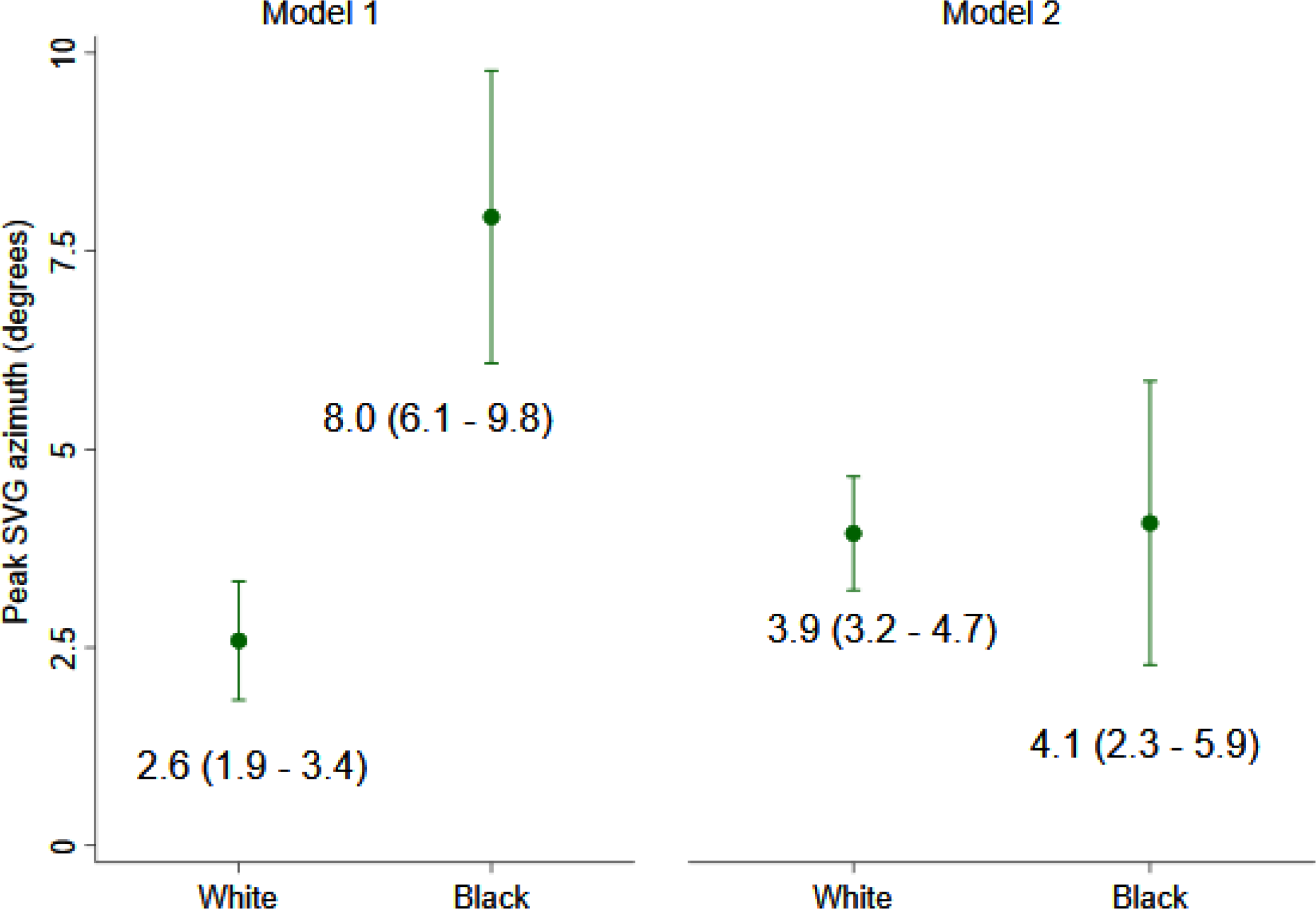
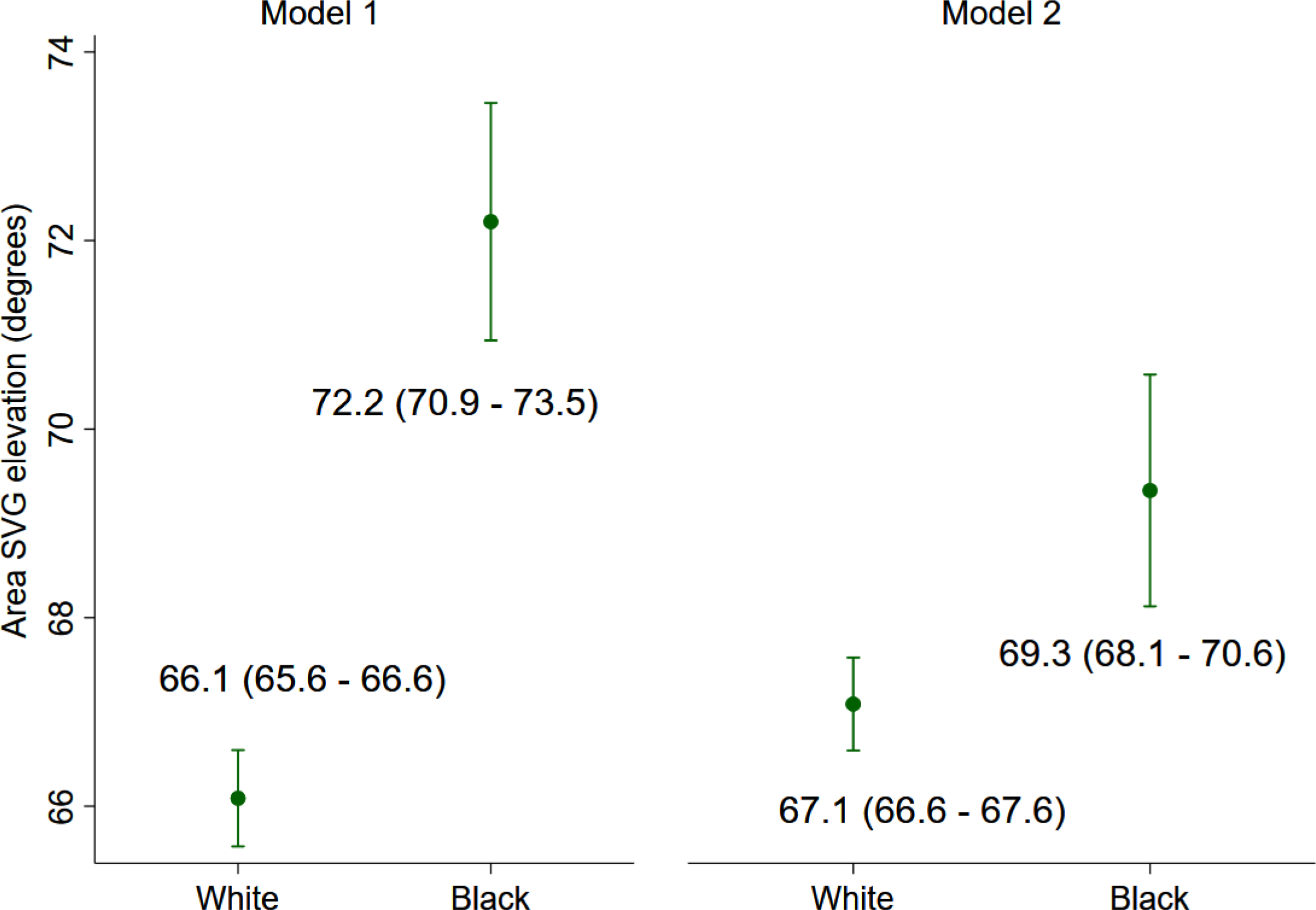
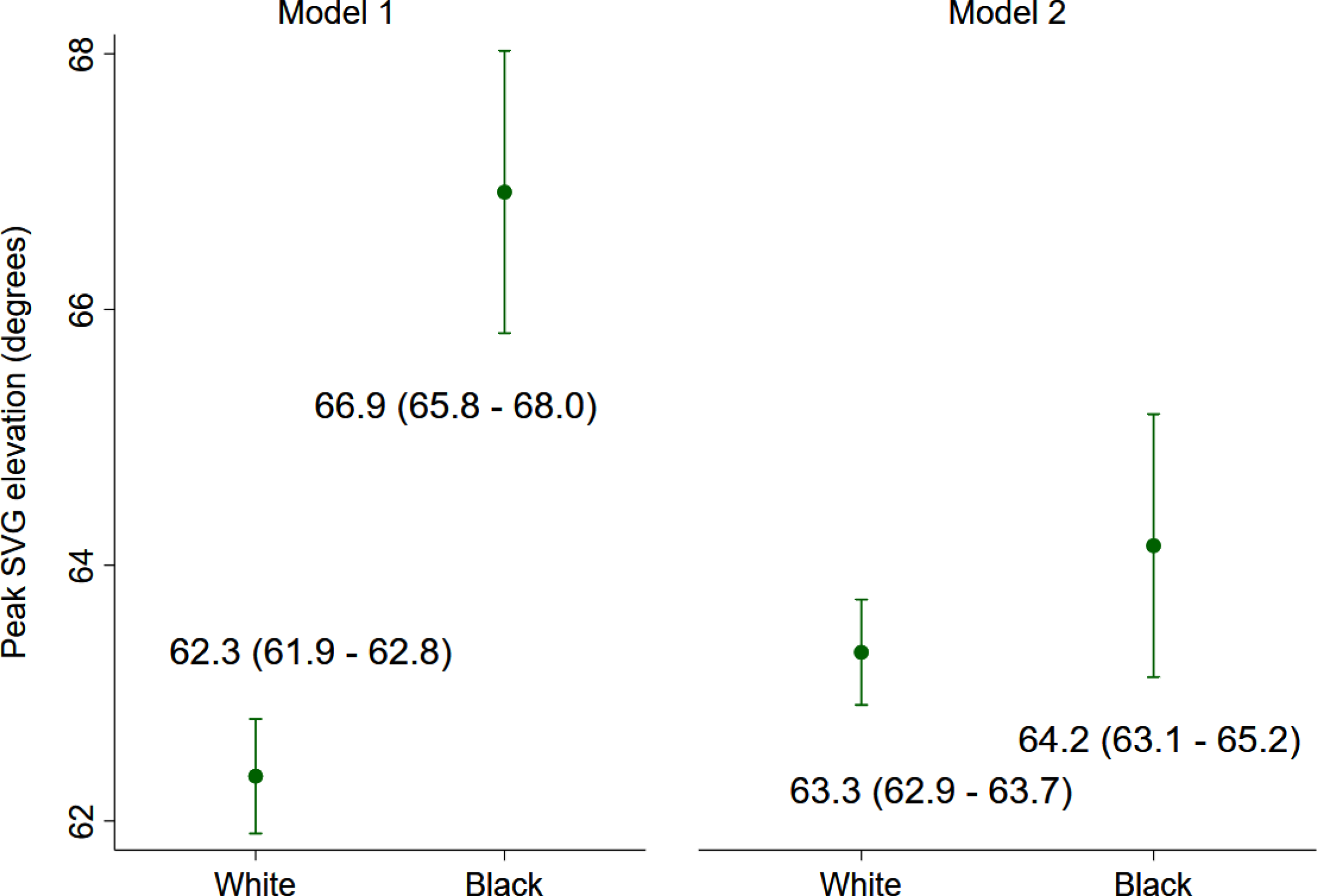
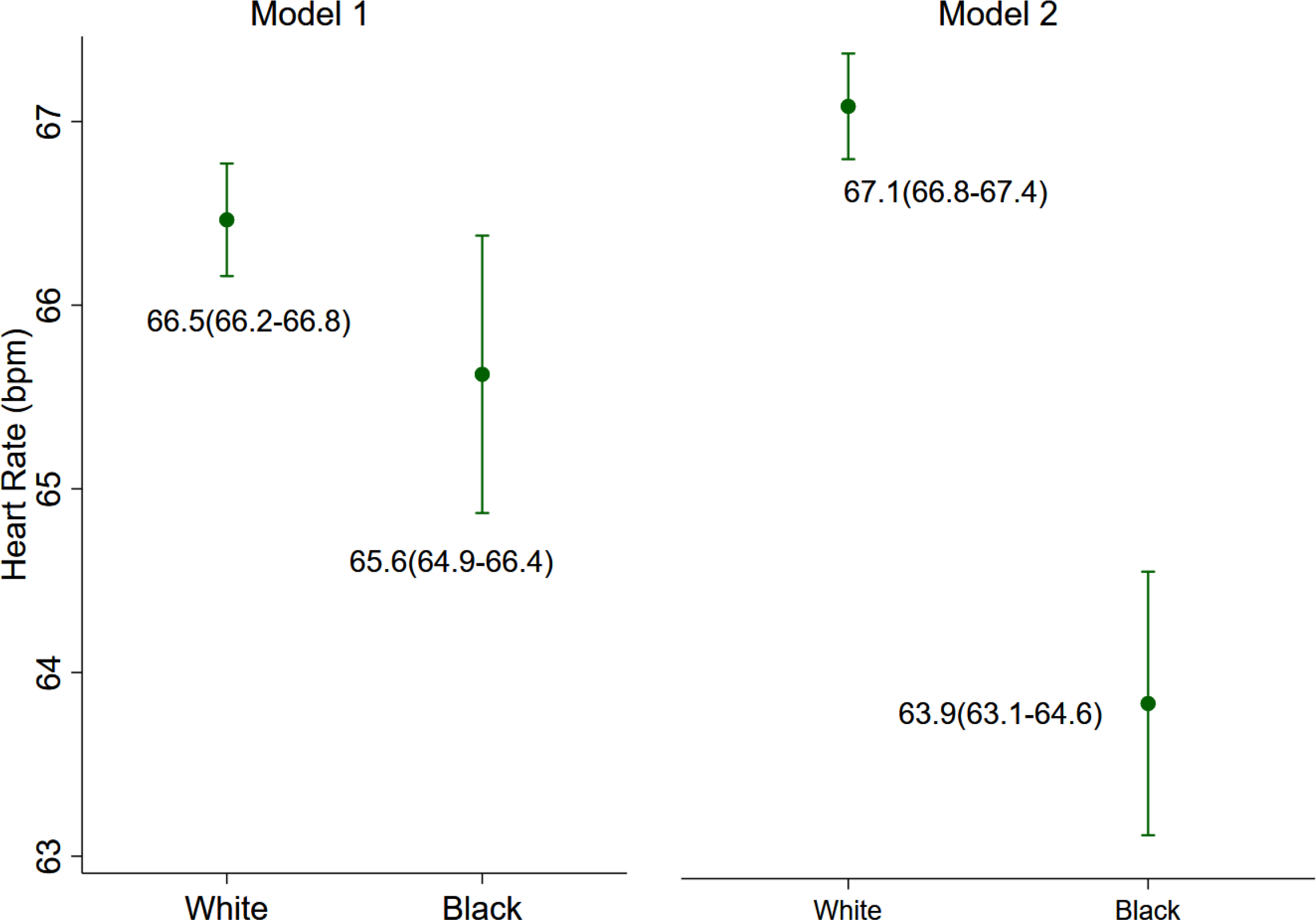
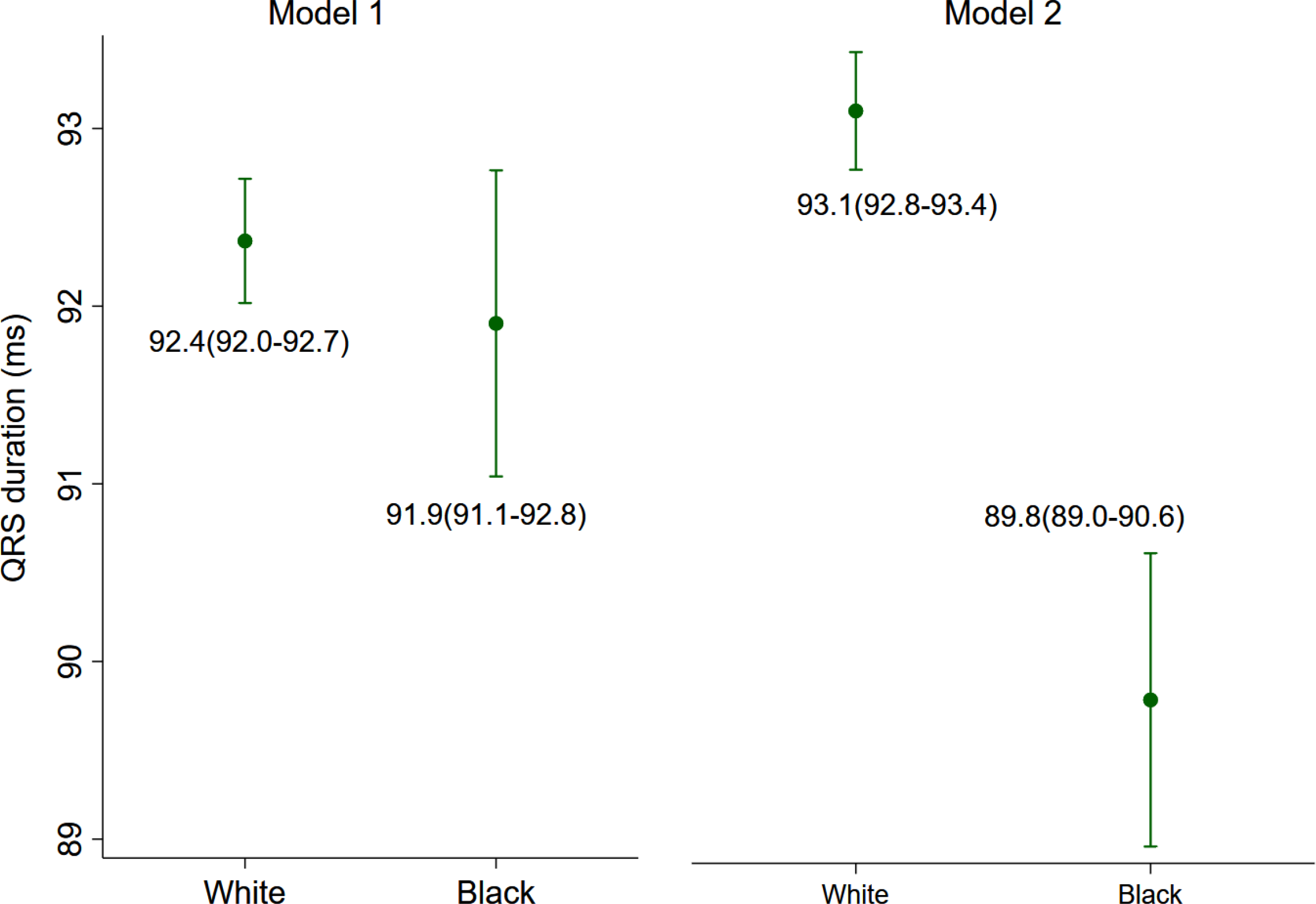
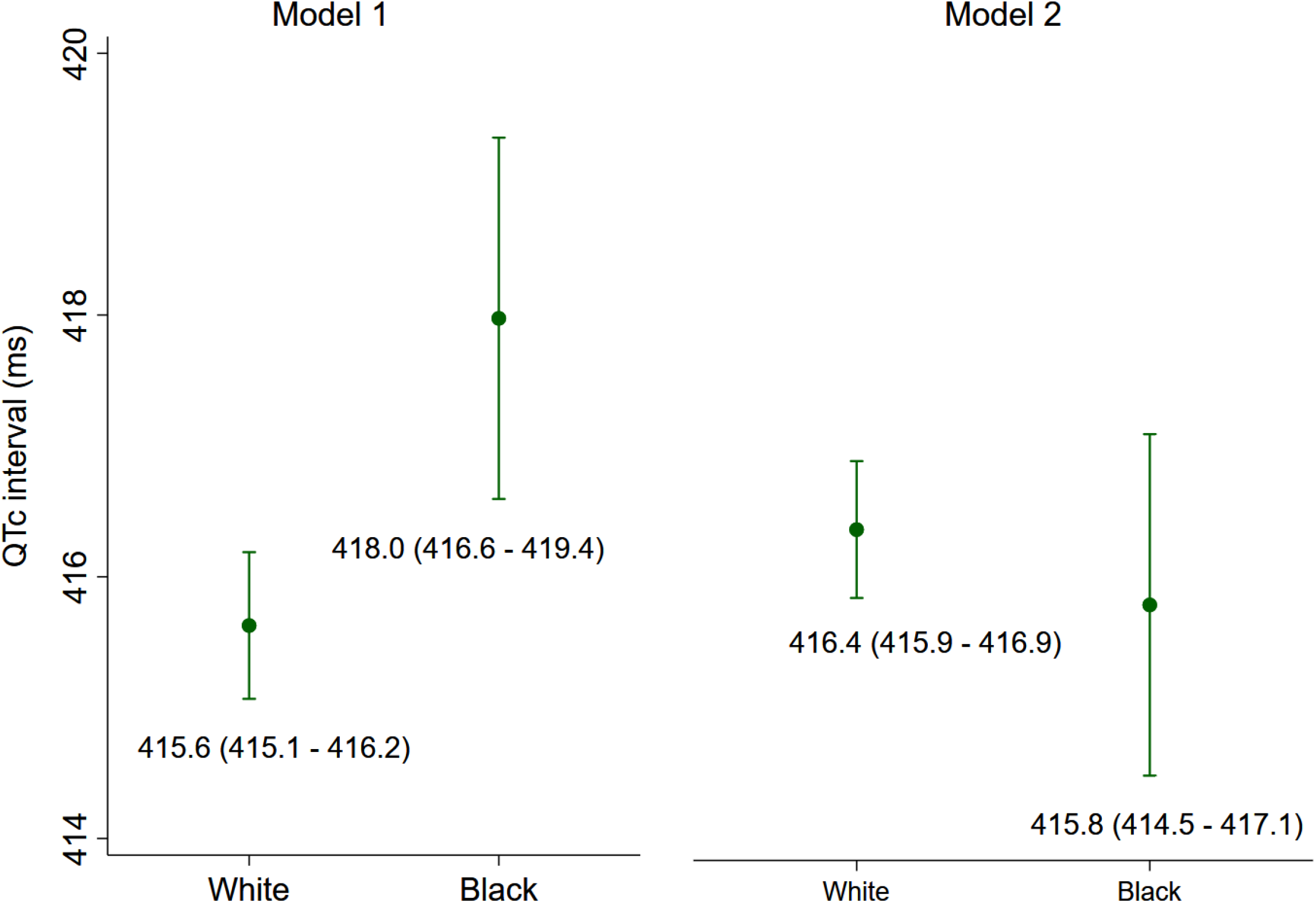
Estimated adjusted marginal (least-squares) means and 95% Confidence Intervals of (**A**) area QRS-T angle, (**B**) peak QRS-T angle, (**C**) area SVG azimuth, (**D**) peak SVG azimuth, (**E**) area SVG elevation, (**F**) peak SVG elevation, (**G**) QRS duration, (**H**) QTc interval for white and black participants. Model 1 was adjusted for age, sex, and study center. Model 2 was additionally adjusted for HF, CHD, stroke, diabetes, hypertension, current smoking and alcohol intake, work, sport, and leisure physical activity levels, levels of total cholesterol, high density lipoprotein, and triglycerides, BMI, use of antihypertensive and antiarrhythmic medications, serum concentrations of sodium, potassium, calcium, magnesium, phosphorus, and uric acid, total protein and albumin, blood urea nitrogen, and CKD stage classified by eGFR_CKD-EPI_, heart rate, QRS duration, Bazett-corrected QT interval, Cornell voltage, education level, occupation category, income, and health insurance.

**Supplemental Figure 2:**
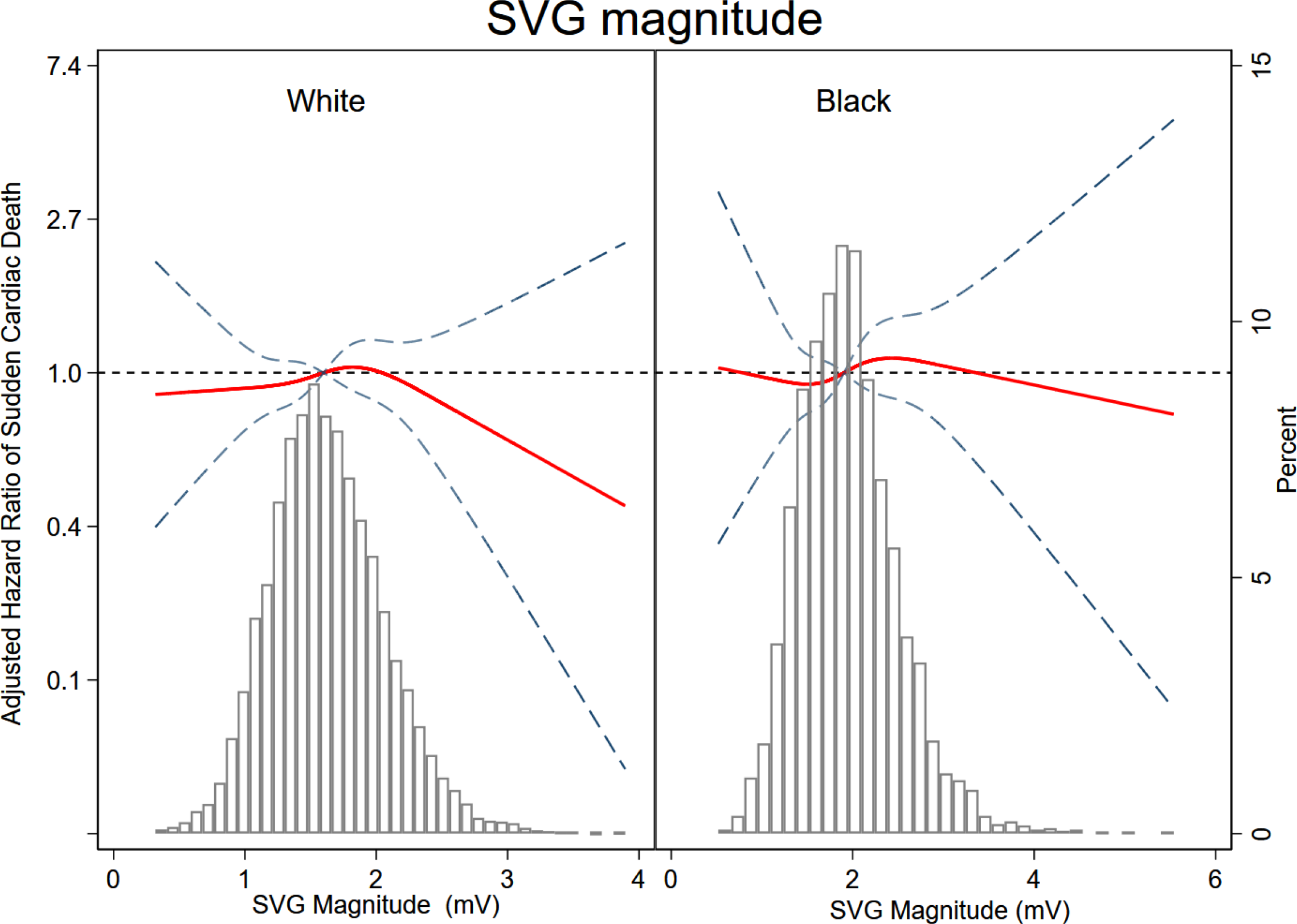
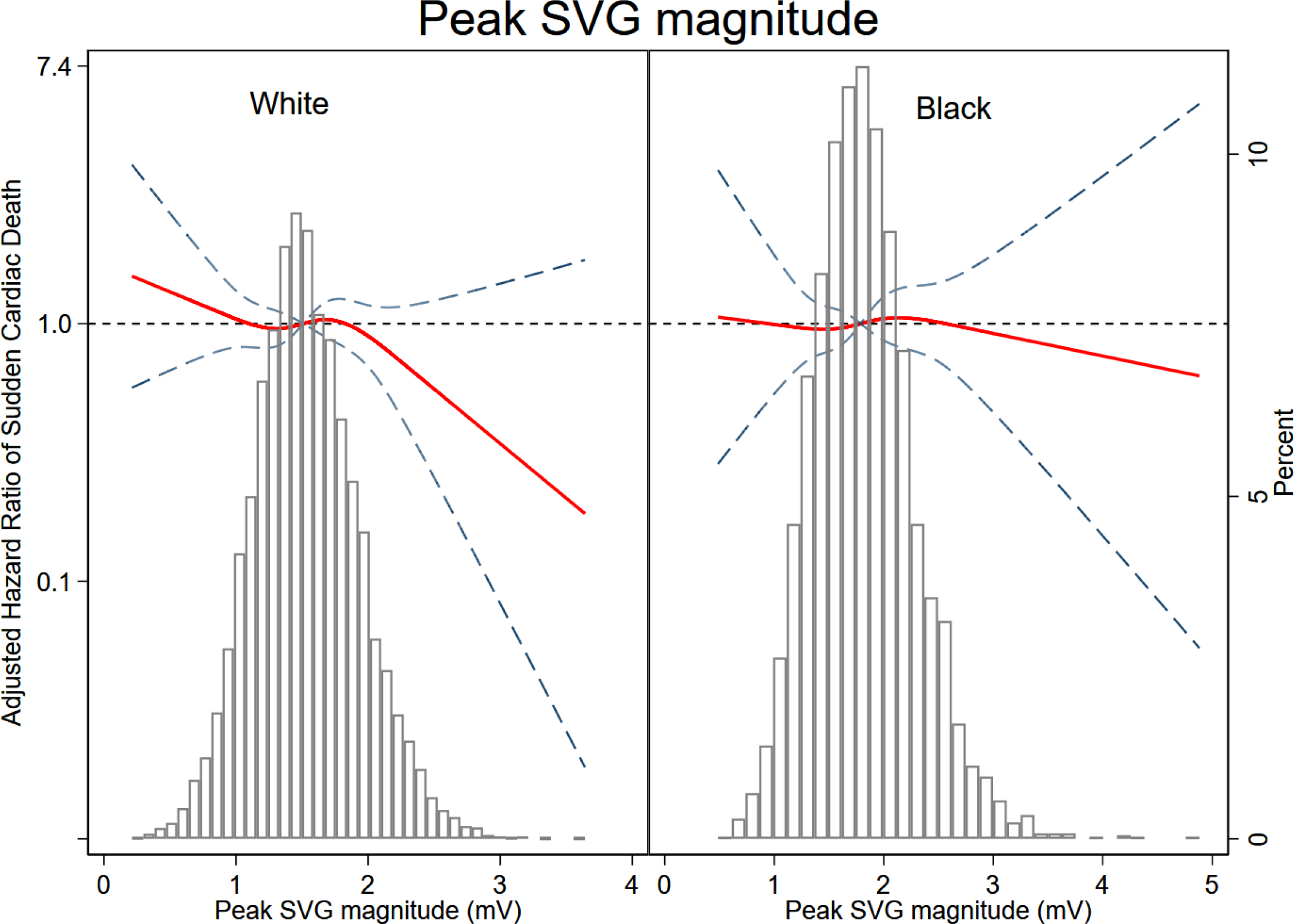
Adjusted (model 1) risk of SCD associated with an area (A) and peak (B) SVG magnitude in black and white participants. Restricted cubic spline with 95% CI shows change in hazard ratio (Y-axis) in response to SVG magnitude change (X-axis). 50th percentile of SVG magnitude is selected as reference. Knots of area SVG magnitude in black participants are at 1.2 – 1.7 – 2.1 – 2.9 mV, and in white participants are at 1.0 – 1.4 – 1.8 – 2.4 mV. Knots of peak SVG magnitude in black participants are at 1.1 – 1.6 – 2.0 – 2.6 mV, and in white participants are at 0.9 – 1.4 – 1.7 – 2.2 mV.

## Notes

### Competing Interest Statement

The authors have declared no competing interest.

### Clinical Trial

not a clinical trial

### Author Declarations

All relevant ethical guidelines have been followed and any necessary IRB and/or ethics committee approvals have been obtained.

Any clinical trials involved have been registered with an ICMJE-approved registry such as ClinicalTrials.gov and the trial ID is included in the manuscript.

